# Independent contributions of language activations in left and right temporal cortex to aphasia outcomes after stroke

**DOI:** 10.1101/2025.09.17.25335931

**Authors:** Sarah M. Schneck, Deborah F. Levy, Jillian L. Entrup, Melodie Yen, Dana K. Eriksson, Marianne Casilio, Anna V. Kasdan, Lily Walljasper, Caitlin F. Onuscheck, Larry T. Davis, Howard S. Kirshner, Michael de Riesthal, Stephen M. Wilson

## Abstract

Recovery from aphasia after stroke has been hypothesized to depend on neuroplasticity in surviving brain regions. Many studies have investigated this process, but progress has been impeded by methodological limitations relating to task performance confounds, contrast validity, and sample sizes. Furthermore, few studies have accounted for the complex relationships that exist between patterns of structural damage, distributed networks of functional activity, and behavioral outcomes.

The present cross-sectional study aimed to overcome these critical methodological limitations and to disentangle the relationships between structure, function, and behavior. We recruited 70 individuals with post-stroke aphasia and 45 neurologically normal comparison participants. We used a valid and reliable language mapping fMRI paradigm that adapted dynamically to each participant’s task performance, and carried out whole-brain permutation analyses along with hypothesis-driven analyses of individually defined functional regions of interest (ROIs). Multivariable models were constructed that incorporated lesion load estimates derived from machine learning and language activations across multiple brain regions.

We found strong evidence that left posterior temporal cortex is the most critical region for language processing in post-stroke aphasia: functional activity in this region was reduced in aphasia, predictive of aphasia outcomes in a whole-brain analysis above and beyond the contribution of lesion load, and remained predictive even above and beyond other functional predictors, with a medium effect size (*f*^2^ = 0.15). We also found that right posterior temporal cortex made an independent contribution to aphasia outcomes: functional activity was attenuated in aphasia, suggesting diaschisis, yet was predictive of aphasia outcomes above and beyond left hemisphere lesion load and functional predictors, with a small effect size (*f*^2^ = 0.08). We corroborated the importance of left frontal cortex: functional activity was attenuated in aphasia and predictive of aphasia outcomes over and beyond the contribution of lesion load; however, unlike in the bilateral temporal regions, functional activity in the left frontal lobe did not remain predictive once other functional predictors were included in the model. There was no support for other potential compensatory mechanisms such as recruitment of the right frontal lobe, the bilateral multiple demand network, or perilesional regions.

Taken together, our findings demonstrate that functional imaging can provide critical insights into language processing in aphasia that cannot be obtained from structural imaging alone, with the left and right posterior temporal cortices making independent contributions to aphasia outcomes after stroke.

## 1 Introduction

Aphasia is an acquired language disorder and is one of the most common consequences of stroke (Pedersen et al., 1995). Fortunately, language function in post-stroke aphasia typically improves over time, though the extent of recovery is highly variable (Kertesz & McCabe, 1977; Wilson et al., 2023). Recovery of language function is thought to reflect neuroplasticity in surviving brain regions, including functional reorganization of language processing and reintegration of spared language areas (Chang & Lambon Ralph, 2020; Hartwigsen & Saur, 2019; Stefaniak et al., 2020). The overarching goal of the present cross-sectional study was to rigorously and comprehensively characterize the neural basis of language processing in individuals with post-stroke aphasia.

Many previous functional imaging studies have investigated the reorganization of language processing in people with aphasia after stroke (Hartwigsen & Saur, 2019). In a recent systematic review and meta-analysis, we showed that there is little consistency in the findings across this literature (Wilson & Schneck, 2021). The most compelling findings attest to the continued importance of residual core left hemisphere fronto-temporal language areas: these regions show reduced functional activity when individuals with aphasia are compared to neurologically normal participants (Crinion & Price, 2005; van Oers et al., 2010) and within cohorts with aphasia, activation of these regions is positively correlated with aphasia outcomes (Crinion et al., 2006; Griffis, Nenert, Allendorfer, Vannest, et al., 2017). There is some evidence that right temporal activation may also be associated with better aphasia outcomes, possibly reflecting pre-existing variability in the capacity of the non-dominant hemisphere (Crinion & Price, 2005).

In contrast, there is limited evidence for other theories of functional reorganization, including compensatory recruitment of right hemisphere homotopic regions (Blank et al., 2003; Leff et al., 2002; Rosen et al., 2000; Tyler et al., 2011; Weiller et al., 1995), bilateral domain-general networks (Brownsett et al., 2014; Geranmayeh et al., 2017; Schneider et al., 2022), or perilesional left hemisphere regions (Fridriksson et al., 2012). We argued that the evidence for these ideas is compromised by three critical methodological limitations. (Wilson & Schneck, 2021). First, task performance confounds are both pervasive and very difficult to control given the inherent difficulty that people with aphasia experience with language tasks (Geranmayeh et al., 2014; Price et al., 2006; Price & Friston, 1999) and can have dramatic effects on functional activation (Binder et al., 2005). Second, many commonly used language contrasts do not actually selectively activate the left-lateralized language network in neurologically normal individuals (Wilson, Yen, et al., 2018) making it challenging to interpret findings in people with aphasia. Third, many studies have included relatively few participants, and perhaps consequently, have often not appropriately corrected for multiple comparisons (Eklund et al., 2016).

Accordingly, the first aim of the present study was to make a concerted effort to overcome these three major methodological limitations (Wilson & Schneck, 2021). To minimize task performance confounds, we used an adaptive language mapping fMRI paradigm in which items were dynamically tailored to real-time participant performance in the scanner (Wilson, Yen, et al., 2018). The validity and reliability of this paradigm has been established by demonstrating that it robustly activates the left-lateralized language network in neurologically normal individuals (Wilson, Yen, et al., 2018). We recruited a large and diverse cohort of 70 individuals with aphasia to maximize statistical power, allowing us to use gold standard whole-brain permutation analyses (Nichols & Holmes, 2002), which we complemented with hypothesis-driven analyses of individually defined functional regions of interest (ROIs).

Another major challenge in the literature to date has been how to address the complex relationships that exist between patterns of structural damage, distributed networks of functional activity in surviving regions, and behavioral outcomes. For example, it could be the case that only individuals with extensive left hemisphere damage have a need for compensatory recruitment of right hemisphere regions, in which case any analysis of right hemisphere functional activity would first need to take into account variability in left hemisphere damage and functional activity (Griffis, Nenert, Allendorfer, Vannest, et al., 2017; Heiss et al., 1999; Heiss & Thiel, 2006; Skipper-Kallal et al., 2017a). The majority of studies have not accounted for structural damage when investigating functional reorganization, despite clear evidence that lesion location and extent are powerful predictors of aphasia outcomes (Kertesz & McCabe, 1977; Levy et al., 2024; Wilson et al., 2023). Moreover, functional reorganization likely relies on the interplay between multiple brain regions (Hartwigsen & Saur, 2019), yet most of the literature involves mass univariate analyses of fMRI data in which functional activity in each voxel is modeled in isolation. There have been some notable exceptions—that is, studies that have investigated the complex relationships between structural damage and functional activity and/or modeled distributed patterns of functional activity (Abel et al., 2015; Fridriksson et al., 2010; Geranmayeh et al., 2016; Griffis et al., 2016; Griffis, Nenert, Allendorfer, & Szaflarski, 2017; Griffis, Nenert, Allendorfer, Vannest, et al., 2017; Johnson et al., 2019; Lorca-Puls et al., 2021; Nenert et al., 2018; Papoutsi et al., 2011; Purcell et al., 2019; Schneider et al., 2022; Sims et al., 2016; Skipper-Kallal et al., 2017a, 2017b; Specht et al., 2009; Stockert et al., 2020; Turkeltaub et al., 2025; Tyler et al., 2010; Warren et al., 2009; Wawrzyniak et al., 2022; Wright et al., 2012)—but although many of these analyses have been nuanced and innovative, they have often been impacted by the three methodological issues outlined above (Wilson & Schneck, 2021).

Consequently, our second aim was to address the complex relationships that hold between lesion location and extent, functional activity in multiple brain regions, and aphasia outcomes, by incorporating structural information and language activations in multiple brain regions into multivariable analyses of functional imaging and behaviour, while maintaining methodological rigor with respect to the critical issues identified above. Specifically, we took the perspective that the most important axis on which individuals with aphasia vary in terms of brain structure is the extent to which their lesions impact the language network (Skipper-Kallal et al., 2017a; Wilson et al., 2023). We therefore used machine learning to empirically determine the extent to which different patterns of structural damage were associated with aphasia outcomes (Levy et al., 2024), which allowed us to derive a scalar lesion load estimate for each individual. We used these lesion load estimates throughout the study to explore how structural variability relates to functional and behavioral observations. Most importantly, we included these lesion load estimates into our functional models to identify brain regions where language activations were predictive of aphasia outcomes *above and beyond* expectations based on lesion location and extent. Finally, we constructed models that incorporated functional activation in multiple brain regions, going beyond the mass univariate approach, to identify which brain regions make unique and independent contributions to aphasia outcomes.

## 2 Methods

### 2.1 Participants

We recruited 70 individuals with chronic or late subacute post-stroke aphasia over a 7-year period (2016−2023). The inclusion criteria were: (1) initial aphasia following left hemisphere supratentorial ischemic or hemorrhagic stroke; (2) at least ∼3 months post-stroke; (3) aged 18 to 90; and (4) fluent and literate in English premorbidly. The exclusion criteria were: (1) neurodegenerative disease, or any other neurological condition impacting language or cognition; (2) significant damage to any right-hemisphere regions homotopic to language regions; (3) major psychiatric disorder; (4) serious substance abuse or withdrawal symptoms; (5) any contraindication to MRI; or (6) compelling evidence of right hemisphere dominance for language. Our study was conducted in accordance with the principles of the 1964 Declaration of Helsinki and was approved by the Institutional Review Boards at Vanderbilt University Medical Center and the University of Arizona.

Of the 70 participants, 48 were recruited acutely at Vanderbilt University Medical Center as part of an ongoing longitudinal study (Wilson et al., 2023) that involves language assessment and neuroimaging at multiple timepoints including ∼3 months and ∼1 year; the latest available timepoint with neuroimaging for each participant was used for the current study. The remaining 22 participants were recruited in the chronic phase of recovery through their participation in aphasia groups or speech-language pathology services in Tucson, Arizona (*n* = 15) (Wilson, Yen, et al., 2018) or Nashville, Tennessee (*n* = 6), or after they had reached out to us with an interest in participating in research (*n* = 1). Approximately 90% of participants had received speech-language treatment; however, treatment intensity, duration, and approach varied considerably across participants.

Forty-five neurologically normal comparison participants were recruited through neighborhood listservs in Nashville, Tennessee (*n* = 32) or Tucson, Arizona (*n* = 13). The inclusion and exclusion criteria were the same as for the individuals with aphasia, except that comparison participants had no neurological history.

Demographic and medical history data are provided in Table 1 and Supplementary Table 1. The comparison participants were approximately matched to the participants with aphasia in terms of age (*P* = .086), sex (*P* = .086), and handedness (*P* = .13), but had completed more education (*P* < .001). To control for potential confounds, these demographic variables were included as covariates in all between-groups analyses.

**Table 1.** Demographic and medical history information.

| | Individuals with aphasia ($n = 70$) | Comparison participants ($n = 45$) |
| --- | --- | --- |
| Age | 59.3 ± 13.4 (32–81) years | 54.5 ± 16.3 (21–83) years |
| Sex | 43 M; 27 F | 20 M; 25 F |
| Handedness | 61 RH; 9 NRH | 34 RH; 11 NRH |
| Education*** | 14.0 ± 3.1 (6–20) years | 16.8 ± 1.9 (12–20) years |
| Lesion extent | 89.7 ± 101.0 (0.7–434.5) cm<sup>3</sup> | |
| Time post onset | 29.8 ± 44.8 (2.8–259.4) months | |
| Stroke type | 63 ischemic, 7 hemorrhagic |  |

Values displayed are mean ± SD (range). M = Male; F = Female; RH = Right-handed; NRH = Non Right-Handed; \*\*\* = Significant group difference at $P < .001$ .

### 2.2 Language evaluation

Language function was evaluated with the Quick Aphasia Battery (QAB), which provides a valid, reliable, time-efficient, and multidimensional characterization of language deficits (Wilson, Eriksson, et al., 2018). The QAB yields eight summary measures, each ranging from 0 (maximal impairment) to 10 (no impairment evident). All analyses in the present study used the QAB overall summary measure, consistent with factor analytic studies showing that a single underlying factor accounts for the majority of variance across different language domains in aphasia (Hula et al., 2010; Kertesz & Phipps, 1977).

All language evaluations were recorded and videotaped for offline scoring. For participants with aphasia, all evaluations were scored by a speech-language pathologist, and most were reviewed in consensus meetings by multiple members of the research team. The 15 individuals with aphasia from Arizona were administered the QAB on three separate occasions (mean 13.2 ± 10.0 days between evaluations), and the summary measures were averaged across administrations. For all but 2 of the remaining participants with aphasia, we administered an extended version of the QAB, which has double the number of word and sentence comprehension items to increase reliability (Wilson et al., 2023).

The 70 participants with aphasia showed substantial variation in both the nature and severity of their language impairments (Table 2; see Supplementary Table 2 for individual QAB scores). At the time of study participation, 15 participants had severe aphasia, 15 moderate aphasia, 19 mild aphasia, and 21 residual aphasia. Most participants had recovered considerably from their initial stroke presentations. Among the 48 participants recruited in the acute phase, 10 were initially untestable but recovered to a QAB overall score of 4.4 ± 3.0 by study participation. The remaining 38 participants recruited acutely recovered from an initial QAB score of 6.2 ± 2.5 to 8.2 ± 2.1 at testing. The 22 participants recruited in the chronic phase had QAB scores of 6.2 ± 2.5 at study participation and had typically recovered from initially moderate or severe aphasia, per their medical histories. This diversity in severity and in the extent of recovery was essential for our correlational analyses, which sought to uncover the neural underpinnings of these disparate outcomes.

**Table 2.** Language evaluation.

|  | Individuals with aphasia | Comparison participants |
| --- | --- | --- |
| Quick Aphasia Battery (QAB) |  |  |
| Word comprehension** | $9.2 \pm 1.6$ (2.9–10.0) | $9.9 \pm 0.2$ (8.8–10.0) |
| Sentence comprehension*** | $6.1 \pm 3.5$ (0.0–10.0) | $9.5 \pm 0.9$ (5.4–10.0) |
| Word finding*** | $6.2 \pm 3.2$ (0.0–10.0) | $9.8 \pm 0.4$ (8.5–10.0) |
| Grammatical construction*** | $6.9 \pm 3.4$ (0.0–10.0) | $9.9 \pm 0.1$ (9.5–10.0) |
| Speech motor programming*** | $7.9 \pm 3.5$ (0.0–10.0) | $10.0 \pm 0.0$ (10.0–10.0) |
| Repetition*** | $6.9 \pm 2.7$ (0.0–10.0) | $9.9 \pm 0.3$ (8.8–10.0) |
| Reading*** | $6.5 \pm 3.3$ (0.0–10.0) | $9.9 \pm 0.3$ (8.8–10.0) |
| QAB Overall*** | $7.0 \pm 2.6$ (0.7–9.9) | $9.8 \pm 0.2$ (9.0–10.0) |
| Written word comprehension | $8.7 \pm 2.6$ (0.0–10.0) | — |
| Pyramids and Palm Trees—Pictures | $13.5 \pm 1.0$ (10–14) | — |

Values displayed are mean ± SD (range). All QAB measures, and the written word comprehension measure are out of 10 points. The short form of Pyramids and Palm Trees is out of 14 points. \*\* = significant group difference at $P < .01$ ; \*\*\* = significant group difference at $P < .001$ .

Written word comprehension and semantic processing, two components of the scanner task, were additionally assessed using items from alternate forms of the QAB and a short version of the Pyramids and Palm Trees—Pictures test (Breining et al., 2015) respectively. These functions were preserved in most, but not all, people with aphasia (Table 2; Supplementary Table 2).

### 2.3 Image acquisition

All participants were scanned on 3 Tesla MRI scanners using standard sequences. Most participants (*n* = 55 with aphasia; *n* = 32 comparison participants) were scanned at the Vanderbilt University Institute for Imaging Science on either a Philips Achieva 3 Tesla scanner or a Philips Ingenia Elition 3 Tesla scanner, each with a 32-channel head coil. T2*-weighted blood oxygen-level dependent (BOLD) echo planar images were acquired with the following parameters: 200 volumes + 4 initial volumes discarded; 35 axial slices in interleaved order; slice thickness = 3.0 mm with 0.5 mm gap; field of view = 220 x 220 mm; matrix = 96 x 96; repetition time (TR) = 2000 ms; echo time (TE) = 30 ms; flip angle = 75 degrees; SENSE factor = 2; voxel size = 2.3 x 2.3 x 3.5 mm. In addition, we acquired high resolution T1-weighted structural images (voxel size = 1.0 x 0.8 x 0.8 mm), FLAIR images to aid in lesion delineation (voxel size = 0.6 x 0.6 x 2.0 mm), and coplanar T2-weighted images to aid in coregistration.

The remaining participants (*n* = 15 with aphasia; *n* = 13 comparison participants) were scanned at the University of Arizona on a Siemens Skyra 3 Tesla scanner with a 20-channel head coil (Wilson, Yen, et al., 2018). T2*-weighted BOLD echo planar images were acquired with the following parameters: 200 volumes + 3 initial volumes discarded; 30 axial slices in interleaved order; slice thickness = 3.5 mm with 0.9 mm gap; field of view = 240 x 218 mm; matrix = 86 x 78; TR = 2,000 ms; TE = 30 ms; flip angle = 90 degrees; voxel size = 2.8 x 2.8 x 4.4 mm. We also acquired T1-weighted structural images (voxel size = 0.9 x 0.9 x 0.9 mm), FLAIR images (voxel size = 0.5 x 0.5 x 2.0 mm), and coplanar T2-weighted images. The 15 individuals with aphasia who were scanned in Arizona completed two functional runs of the language mapping paradigm 10.3 ± 7.0 days apart.

Each scanner had a button box to collect participant responses, and a screen located behind the bore to display visual stimuli, which participants viewed through a mirror attached to the head coil.

### 2.4 Structural imaging

#### 2.4.1 Lesion delineation and image processing

Lesions were manually delineated using ITK-SNAP version 3.6.0 (Yushkevich et al., 2006), making reference to T1-weighted and FLAIR images. The T1-weighted images were warped to Montreal Neurological Institute (MNI) space. To reduce the impact of lesions on normalization in participants with aphasia, the lesion was replaced prior to normalization with healthy tissue from the opposite hemisphere—enantiomorphic normalization (Nachev et al., 2008)—implemented with in-house code in Matlab R2020a. Then, the images were normalized using Unified Segmentation (Ashburner & Friston, 2005) implemented in SPM12. Infarcts varied widely in location and extent, but were mostly situated in middle cerebral artery (MCA) territory (Table 1; Fig. 1A).

**Figure 1.**
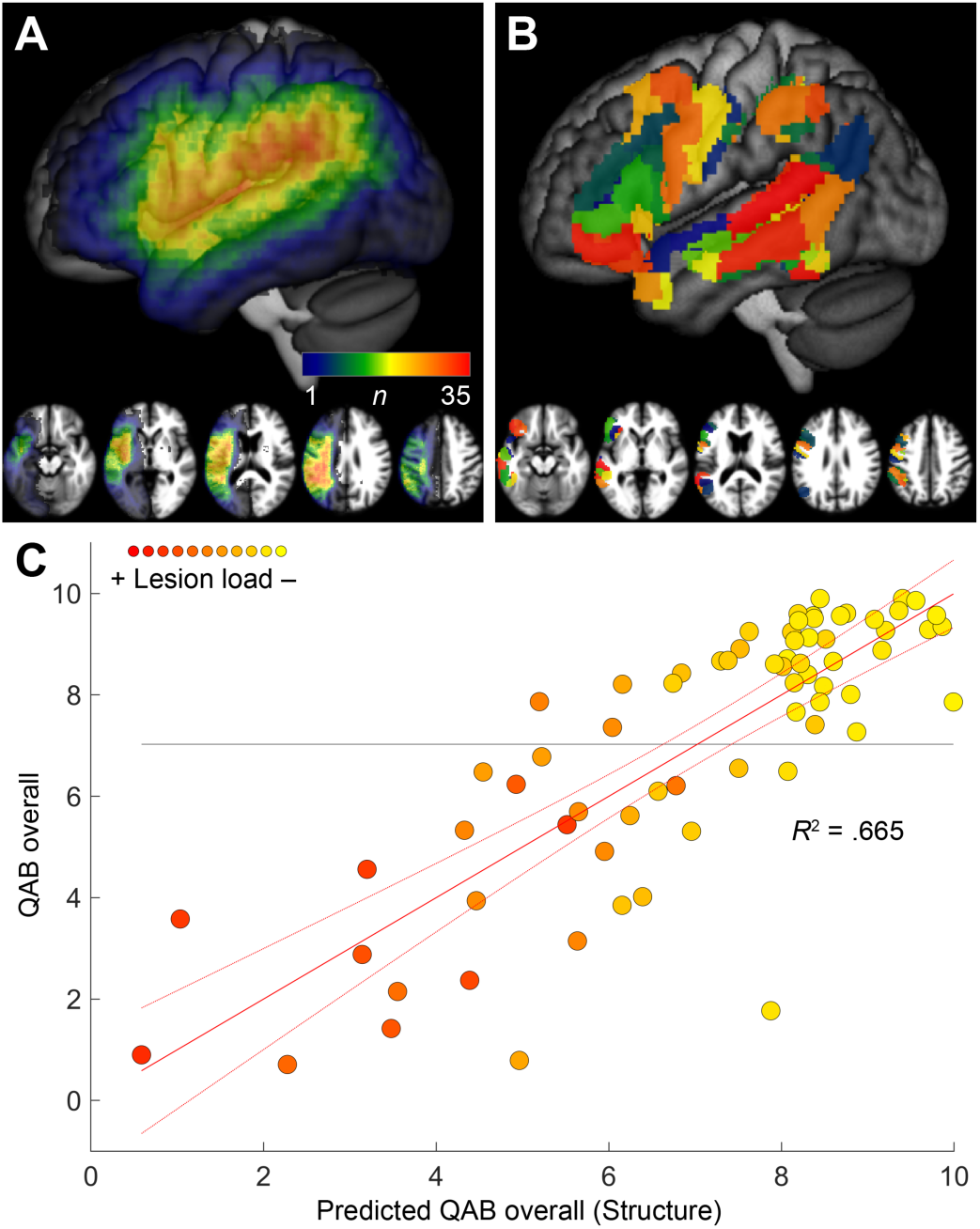
Analysis of structural imaging data. (**A**) Lesion overlay. *n* = Number of people with aphasia with damage to each voxel. (**B**) Regions of interest (ROIs) from the human Brainnetome Atlas used for predicting QAB overall from structural data. (**C**) Prediction of QAB overall from structural data (i.e., lesion masks) and covariates (age, sex, handedness, education, stroke type, time post onset). The red solid line depicts the line of best fit, the red dotted lines indicate the 95% confidence interval of the fit, and the horizontal line depicts the mean observed QAB overall score. Each data point is color-coded to indicate the individual’s lesion load (i.e., predicted QAB overall score based on structural imaging only) on a scale from yellow (less lesion load) to red (greater lesion load), in this and subsequent figures.

#### 2.4.2 Lesion load proxy

We sought to quantify each individual’s lesion load with a single scalar measure with which we could (1) depict this fundamental dimension of variability throughout our study and (2) incorporate the impact of structural damage into our functional analyses. We operationalized lesion load by using support vector regression to empirically determine the extent to which different patterns of structural damage were associated with aphasia outcomes (QAB overall scores), similar to the approach we have described previously (Levy et al., 2024). Specifically, we parcellated the left hemisphere language network into 44 spatial ROIs, and created a 44-dimensional vector for each individual with aphasia, encoding the lesion impact in each spatial ROI. The 44 ROIs are displayed in Fig. 1B and were identified based on overlap (> 20 voxels, 160 mm^3^) between the human Brainnetome Atlas (Fan et al., 2016) and a functionally defined language network. The human Brainnetome Atlas defines 246 left hemisphere regions with sufficient granularity to distinguish adjacent regions, that may have distinct functions in language processing. The functional language network was defined as lateral frontal, temporal, or parietal regions that were activated for semantic or phonological matching tasks in a different group of neurologically normal individuals (Yen et al., 2019) (*n* = 16, cluster-defining threshold: *P* < .005; corrected at *P* < .05 based on cluster extent using permutation analysis); these activated regions were dilated by approximately 6 mm. To calculate the lesion impact in each ROI, we intersected the 44 spatial ROIs with individual lesion masks that were smoothed with an 8 mm full width at half maximum (FWHM) Gaussian kernel. Finally, a QAB overall score was predicted for each participant with aphasia using support vector regression with a linear kernel and box constraint *C* = 1, implemented with the *fitsvm* and *kfoldPredict* functions in Matlab R2020a. A leave-one-out cross-validation procedure was used, such that each participant’s predicted QAB overall score was calculated based on the relationships between lesion impact vectors and observed QAB overall scores in the remaining 69 participants with aphasia. These predicted QAB overall scores represent the expected impact of each individual’s lesion on language function and were thus subsequently used as scalar representations of lesion load. The relationship between predicted and observed QAB overall scores is shown in Fig. 1C. As can be seen, aphasia outcomes were predicted quite accurately based on structural imaging. The three ROIs where damage was most predictive of poorer outcomes were all localized to the posterior temporo-parietal region. Previous studies have shown that aphasia can be predicted more accurately based on information about lesion location than lesion extent alone (Hope et al., 2013; Levy et al., 2024). Lesion load estimates were used to characterize the variability in language network integrity in our aphasia cohort in visual depictions of analyses. Specifically, in all figures where data from individual participants with aphasia are displayed, each data point is color-coded (on a scale from yellow to red) to indicate the individual’s lesion load (i.e., predicted QAB overall score based on structural imaging only).

### 2.5 Functional imaging

#### 2.5.1 Language mapping paradigm

To identify brain regions involved in language processing, we used an Adaptive Language Mapping (ALM) paradigm that we developed previously (Wilson, Yen, et al., 2018). This paradigm involved an AB block design in which a semantic matching language condition was contrasted with a perceptual matching control condition (20-s blocks; 10 blocks per condition; 6:40 total scan time). These two conditions were largely matched for visual, motoric, and cognitive demands but critically differed in terms of language processing. In the semantic matching condition, participants viewed two written words, one above the other, and decided if the words “go together”, i.e., if they were semantically related (Fig. 2A). In the perceptual matching condition, participants viewed two false font strings, one above the other, and decided if the strings were identical (Fig. 2B). In both conditions, participants pressed a button with their left hand for any trial they perceived to be a match (for non-matches, they were instructed to do nothing).

**Figure 2.**
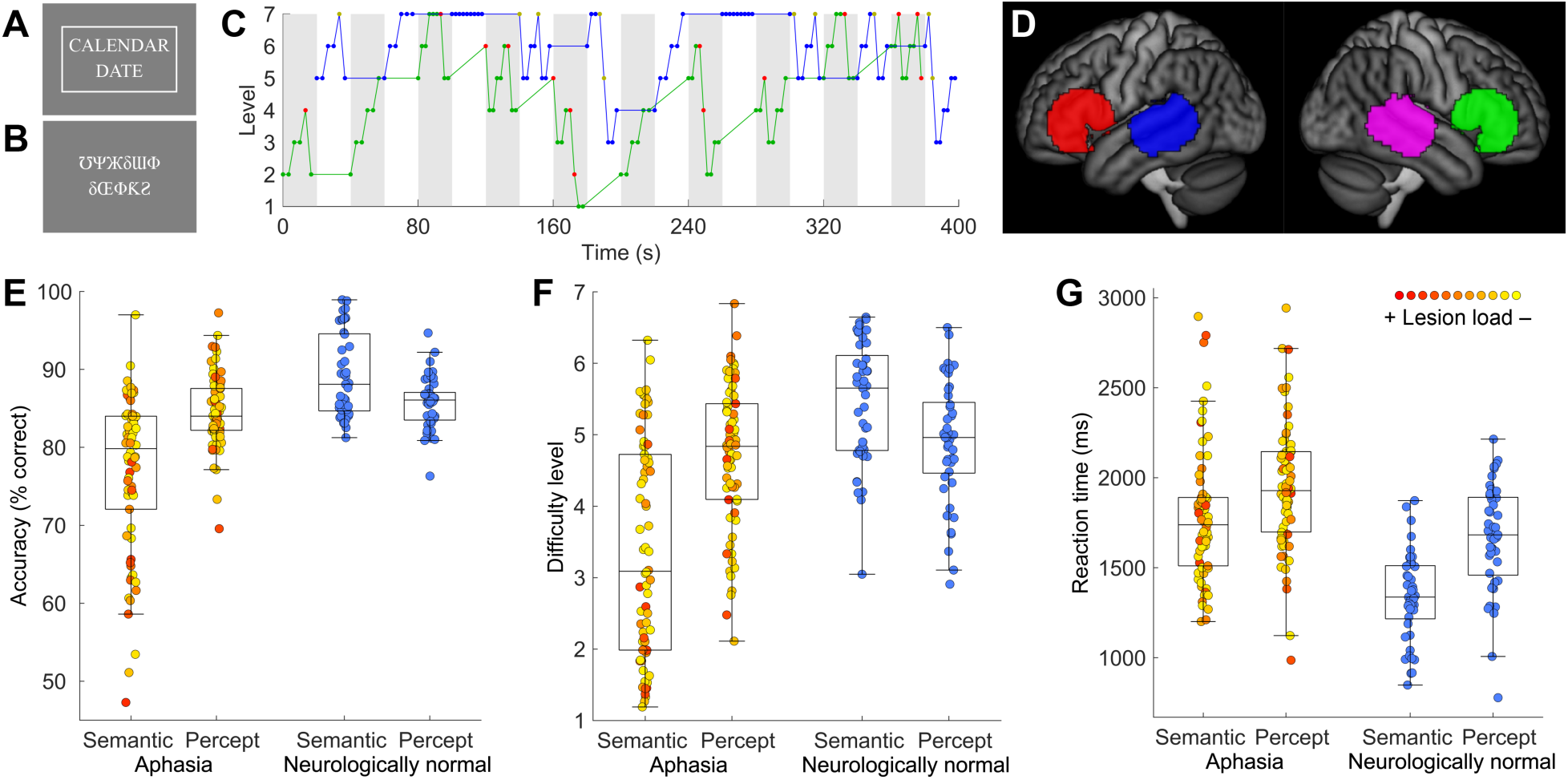
Functional MRI paradigm and behavioral data. (**A**) Example item from the Semantic condition. In this condition, participants viewed two written words, one above the other, and decided if the words “go together”, i.e., if they were semantically related. The rectangle around the words indicates that the ‘match’ button has been pressed. (**B**) Example item from the Perceptual condition. In this condition, participants viewed two false font strings, one above the other, and decided if the strings were identical. This is a mismatch trial, so the ‘match’ button has not been pressed. (**C**) The adaptive staircase procedure in a representative participant. We used a 2-down-1-up adaptive staircase procedure with weighted step sizes and 7 levels of difficulty, which aimed to keep all participants at around 80% accuracy. Grey = Semantic condition; White = Perceptual condition; Green line = Semantic level of difficulty; Green dots = Correct Semantic trials; Red dots = Error Semantic trials; Blue line = Perceptual level of difficulty; Blue dots = Correct Perceptual trials; Yellow dots = Error Perceptual trials. (**D**) Search regions for functional regions of interest (ROIs) that were defined in individual participants. (**E**) Accuracy on the scanner task in individuals with aphasia and neurologically normal participants. (**F**) Difficulty level. (**G**) Reaction time. The color of each aphasia datapoint indicates the lesion load estimate.

Critically, both conditions independently adapted to each individual’s real-time performance, such that as an individual responded correctly, the condition became more difficult, and as an individual responded incorrectly, the condition became easier. Specifically, a 2-down-1-up adaptive staircase procedure with weighted step sizes and 7 levels of difficulty was used (Fig. 2C). After two consecutive correct responses, the level of difficulty increased by one level on the subsequent trial, and after any incorrect response, the difficulty was decreased by two levels on the subsequent trial. This procedure aimed to keep all participants at around 80% accuracy (García-Pérez, 1998). The degree of difficulty in the semantic condition was manipulated by modulating the frequency, concreteness, length, and age of acquisition of the two words; the closeness of the match; and the presentation rate, which ranged from 4 to 10 items per block. The degree of difficulty for the perceptual control condition was manipulated by modulating the similarity of the mismatching pairs and the presentation rate, which also ranged from 4 to 10 items per block.

The paradigm was implemented using Matlab (Mathworks, Natick, MA) and the Psychophysics Toolbox (Kleiner et al., 2007). All participants were trained on the paradigm before entering the scanner and had a further opportunity to practice inside the scanner while initial structural scans were being obtained.

The ALM paradigm was developed to address two specific methodological challenges. The first challenge involves task performance confounds. Individuals with aphasia inherently find language tasks much more difficult than do those without aphasia, which means that comparisons between people with aphasia and neurologically normal participants, and comparisons between people with aphasia varying in severity, are confounded by task difficulty. Therefore, it can be hard to determine whether any observed activation differences reflect functional reorganization as opposed to effects of accuracy or reaction time (Binder et al., 2005; Fridriksson & Morrow, 2005; Geranmayeh et al., 2014; Price et al., 2006; Price & Friston, 1999). In our analysis of the broader literature, we found that the majority of contrasts and second level analyses failed to match accuracy, reaction time, or cognitive demands in general, and in many cases individuals with aphasia were unable to perform language tasks in the scanner, or there was insufficient data reported to establish whether they could perform language tasks (Wilson & Schneck, 2021). The ALM paradigm was designed to confront this problem by dynamically selecting items according to each individual’s real-time performance, with the goal of presenting each participant, regardless of language abilities, with tasks in the scanner that are difficult but achievable (Wilson et al., 2019; Wilson, Yen, et al., 2018).

The second challenge is that many commonly used language contrasts do not selectively activate the left-lateralized language network in neurologically normal individuals (Wilson, Yen, et al., 2018; Wilson & Schneck, 2021). This is a major limitation, because aphasia typically results from left hemisphere damage, and so fMRI paradigms that selectively activate left hemisphere language regions (in undamaged brains) are a prerequisite for understanding how these core language functions might be reorganized after the left hemisphere language network is damaged (Wilson & Schneck, 2021). The ALM paradigm has been validated, and empirically shown to reliably activate left-lateralized language regions in neurologically normal individuals, making it an ideal tool for investigating potential reorganization of language function (Wilson, Yen, et al., 2018). Note that although ALM is a word-level semantic decision paradigm, the core left hemisphere language regions that it reveals are essentially the same as those seen in sentence or discourse level paradigms (Diachek et al., 2022; Wilson, Yen, et al., 2018). Our rationale for using ALM does not relate to the specific language task or subtraction it employs, but rather its demonstrated ability to reliably map core left hemisphere language areas.

#### 2.5.2 Image processing and model fitting

The functional imaging data were first preprocessed with tools from AFNI (Cox, 1996). Slice timing and head motion were corrected, with six translation and rotation parameters saved for use as covariates. Next, the data were proportionally scaled to a global mean of 10,000 within a brain mask to account for inter-scanner intensity differences, then the data were smoothed with a Gaussian kernel (FWHM = 6 mm) and detrended with a Legendre polynomial of degree 2. Finally, independent component analysis was performed using the FSL tool *melodic* (Beckmann & Smith, 2004). Noise components were manually identified (Kelly et al., 2010) and removed using *fsl_regfilt*.

The ALM paradigm was modeled with a boxcar function convolved with a standard hemodynamic response function and fit to the data using *fmrilm* from the FMRISTAT package (Worsley et al., 2002). Language maps were constructed by contrasting the semantic matching and perceptual matching conditions. All voxels were included in whole-brain analyses, thus absent or attenuated activation may reflect structural damage or functional changes. For the 15 individuals with aphasia from Arizona who had two functional runs, the two runs were combined with fixed effects models. Activation maps were coregistered to coplanar T2-weighted images, then to T1-weighted structural images, and then normalized to MNI space.

#### 2.5.3 Individually defined functional ROIs

In addition to whole-brain voxelwise analyses, we carried out a set of parallel analyses based on individually defined functional ROIs (Fedorenko et al., 2010). This approach served two purposes. First, the use of a small number of hypothesis-driven ROIs improved sensitivity by allowing us to conduct analyses without needing to correct for multiple comparisons at the whole-brain level. Second, the use of individual functionally defined ROIs permitted the quantification of activations for language processing in spatial locations that could differ across participants (Fedorenko et al., 2010). This may be especially beneficial in stroke survivors, in whom it is plausible that language activations may be displaced from typical locations, due to the impact of lesions and potential functional reorganization (Blank et al., 2017)

We followed our methods described previously (Quillen et al., 2021) loosely based on earlier work (Fedorenko et al., 2010). Four large search space ROIs were defined as follows. First, language activation peaks were identified in the left temporal lobe and left frontal lobe in a separate group of 20 previously scanned neurologically normal participants performing a semantic matching task (Quillen et al., 2021). The left temporal peak was located at MNI coordinates (−56, −34, 0) and the left frontal peak at (−50, 24, 2). We included just these two core language regions because a more comprehensive parcellation of the language network would require a larger study, given the similarity of functional response profiles across the network (Blank et al., 2016). Next, these two peaks were mirrored to the right hemisphere. Then, we identified voxels within 24 mm of each peak or mirrored peak, subject to the requirement that temporal ROIs overlapped any human Brainnetome Atlas ROIs in the superior temporal gyrus (STG), superior temporal sulcus (STS), middle temporal gyrus (MTG), or inferior temporal gyrus (ITG), and frontal ROIs overlapped any atlas ROI in the inferior frontal gyrus (IFG), middle frontal gyrus (MFG), superior frontal gyrus (SFG), precentral gyrus, orbital gyrus, or paracentral lobule. This ensured that search space ROIs did not extend into other lobes or white matter. The four search space ROIs are shown in Fig. 2D.

To quantify language responses in individually specific language voxels within these large search space ROIs, we next fit new models to each participant’s functional data, modeling odd-and even-numbered blocks separately, in order to partition the data into localizer and response testing components. The voxels with the top 20% of *t* statistics for the odd-numbered blocks within each search space ROI were identified (subject also to *t* > 0). Then, the signal change estimates for these voxels from the even-numbered blocks (i.e., the blocks not used to select the voxels) were recorded and averaged across voxels. This procedure was repeated, using the even-numbered blocks to select voxels and the odd-numbered blocks to obtain effect sizes. Finally, the two effect size estimates (odd-to-even and even-to-odd) were averaged together. The upshot of this procedure is that it quantifies language activations that are replicable (because separate data are used to identify voxels and quantify responses) yet can be individually localized anywhere within the large search space ROIs.

The mean proportion of the ROIs included in lesion masks was 22% ± 31% (range 0−100%) for the left temporal ROI and similarly 22% ± 32% (range 0−100%) for the left frontal ROI. All voxels were considered within the search spaces, regardless of structural damage; note that in the presence of substantial damage or lack of actual activation, this approach will yield signal estimates around zero because noise will not replicate across odd and even blocks. For the 15 individuals with aphasia from Arizona who had two functional runs, the procedure described was carried out separately for each run, then effect size estimates were averaged across the two runs.

### 2.6 Statistical analysis

The behavioral data from task performance in the scanner were analyzed with repeated measures general linear models using *fitlm* in Matlab R2020a. There was a between-subjects fixed effect of group (individuals with aphasia; comparison participants); a within-subjects fixed effect of condition (semantic matching; perceptual matching); and covariates of age, sex, handedness (right-handed versus non-right-handed, i.e., left-handed or ambidextrous), and education (years).

To analyze the fMRI data, we carried out a series of whole-brain voxelwise analyses, and a parallel series of analyses based on individually defined functional ROIs (Fedorenko et al., 2010). The whole-brain analyses were corrected for multiple comparisons using permutation testing (thresholded based on cluster extent at *P* < .05; cluster-defining threshold: *P* < .005; 1000 permutations; implemented with *randomise_parallel*). The ROI analyses were performed with *fitlm* in Matlab R2020a and each set of four models (one for each ROI) was corrected for multiple comparisons using the false discovery rate (FDR) procedure (Benjamini & Hochberg, 1995).

We first mapped language processing in individuals with aphasia, and separately in comparison participants using 1-sample voxelwise *t*-tests, then we compared these two groups directly in whole-brain and ROI analyses, including covariates of age, sex, handedness, and education.

Second, to identify brain regions where activation for language processing was correlated with aphasia outcome (QAB overall) in individuals with aphasia, we carried out whole-brain and ROI analyses with QAB overall score as the dependent variable. In addition to the covariates just described, these analyses included covariates of stroke type (ischemic, hemorrhagic) and time post onset (logarithmically transformed).

Third, we identified brain regions where activation for language processing was predictive of aphasia outcome (QAB overall) above and beyond expectations based on structural damage (i.e., the lesion load variable, which was operationalized as the predicted QAB overall score based on structural damage). Again, whole-brain and ROI analyses were performed. Aphasia outcome (QAB overall) was again the dependent variable but, critically, the lesion load variable was also included in the models (along with the covariates just described). This required any significant finding to be significant *above and beyond* what structural information was able to explain. We also included the interaction of lesion load by time post onset, which accounted for significant variance because our more chronic participants tended to have greater lesion loads, since they had been mostly recruited in the context of long-term involvement in aphasia groups.

Finally, we included language activations from multiple functional ROIs simultaneously in a single model, along with the lesion load variable and the covariates, to investigate which ROIs made unique and independent contributions to accounting for aphasia outcomes. In other words, this final analysis sought to identify brain regions in which language activations were predictive of aphasia outcomes not only above and beyond what structural information could explain, but also above and beyond what language activations in other brain regions could explain.

## 3 Results

### 3.1 Functional MRI task performance

In the scanner, 67 out of 70 individuals with aphasia performed above chance on the language condition of the ALM paradigm, and all 70 performed above chance on the perceptual condition, confirming task compliance (Fig. 2E,F,G). There were, however, significant interactions of group by condition for accuracy (*F*(1, 109) = 25.019; *P* < .001), mean difficulty level (*F*(1, 109) = 28.71; *P* < .001), and reaction time (*F*(1, 109) = 6.24; *P* = .014), indicating that the aphasia group was disproportionately impaired on the language condition.

For accuracy, the aphasia group was less accurate on the semantic condition relative to the perceptual condition (semantic accuracy: 76.8% ± 10.1%; perceptual accuracy: 84.6% ± 4.6%; *t*(69) = −5.70, *P* < .001) while the comparison group showed the opposite pattern (semantic accuracy: 89.2% ± 5.3%; perceptual accuracy: 85.7% ± 3.3%); *t*(44) = 4.03, *P* < .001). In the aphasia group, QAB overall scores (indicating overall language processing function) were correlated with accuracy on the semantic condition (*r* = .57, *P* < .001), but not on the perceptual condition (*r* = −.02, *P* = .90). Note that the 3 participants who performed at chance on the language condition all showed evidence of deficits on the task components of written word comprehension and semantic processing (see dataset on OSF for details).

For mean difficulty level, the aphasia group performed at lower levels on language items than perceptual items (semantic mean level: 3.4 ± 1.5; perceptual mean level: 4.7 ± 1.0; *t*(69) = −6.09, *P* < .001) while the comparison group showed the opposite pattern of performance (semantic mean level: 5.4 ± 0.8; perceptual mean level: 4.9 ± 0.8; *t*(44) = 3.88, *P* < .001). In the aphasia group, QAB overall scores were correlated with the mean difficulty level of the semantic condition (*r* = .43, *P* < .001), but not of the perceptual condition (*r* = .05, *P* = .68).

For reaction time, both groups responded more quickly on the semantic condition, but the aphasia group showed a smaller advantage for the semantic condition (semantic reaction time: 1780 ms ± 368 ms; perceptual reaction time: 1949 ms ± 354 ms; *t*(69) = −4.25, *P* < .001) than did the comparison group (semantic reaction time: 1339 ms ± 241 ms; perceptual reaction time: 1664 ms ± 298 ms; *t*(44) = −10.89, *P* < .001). In the aphasia group, QAB overall scores were negatively correlated with reaction times on the semantic condition (*r* = −.29, *P* = .016), but not on the perceptual condition (*r* = .00, *P* = .99).

### 3.2 Functional MRI task activation patterns

The aphasia group activated a strongly left-lateralized network for the contrast of the semantic condition with the perceptual condition (Fig. 3A; Table 3). Activated regions included the whole length of the left STS extending ventrally to the posterior MTG and posterior ITG, and posteriorly to the angular gyrus; the left IFG (pars opercularis, triangularis, and orbitalis) extending posteriorly to the dorsal and ventral precentral gyrus; the right IFG (pars triangularis and opercularis); and the right cerebellum. Comparison participants activated a similar network (Fig. 3B; Table 3).

**Figure 3.**
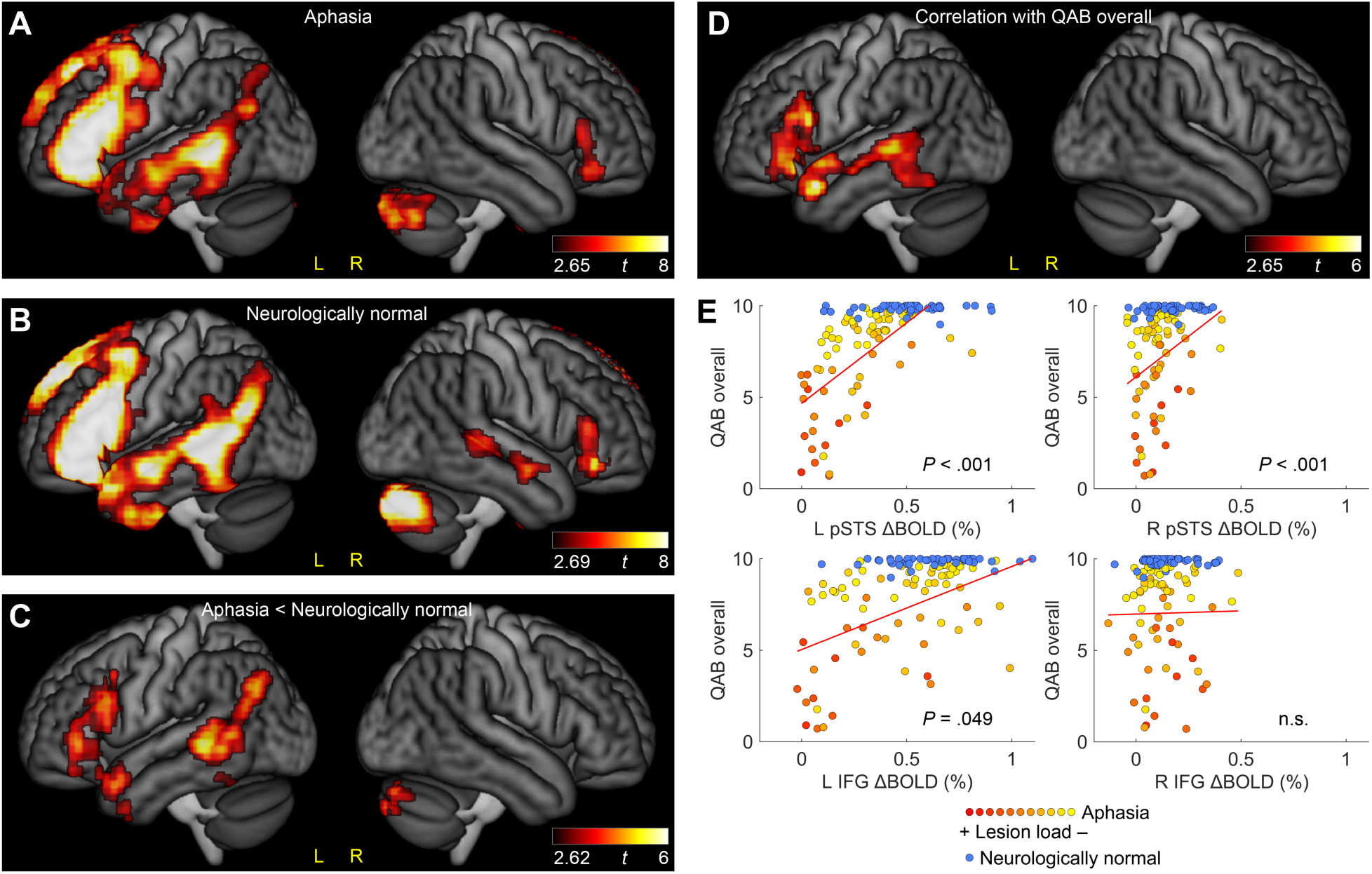
Standard functional MRI analysis. (**A**) Brain regions activated for the Semantic condition compared to the Perceptual condition in 70 individuals with aphasia. (**B**) Brain regions activated for the Semantic condition compared to the Perceptual condition in 45 neurologically normal comparison participants. (**C**) Brain regions that were less activated for language processing in individuals with aphasia compared to neurologically normal participants. Note that there were no regions that were more activated in the aphasia cohort. (**D**) Brain regions where BOLD signal change was correlated with Quick Aphasia Battery (QAB) overall scores. (**E**) The association between BOLD signal change in individually defined regions of interest (ROIs) and QAB overall scores in individuals with aphasia (hot colored datapoints) and neurologically normal comparison participants (blue datapoints). The color of each aphasia datapoint indicates the lesion load estimate. The red fit lines are based only on the aphasia cohort. These plots are not adjusted for covariates (but covariates were included in the analyses described in the main text).

**Table 3.**
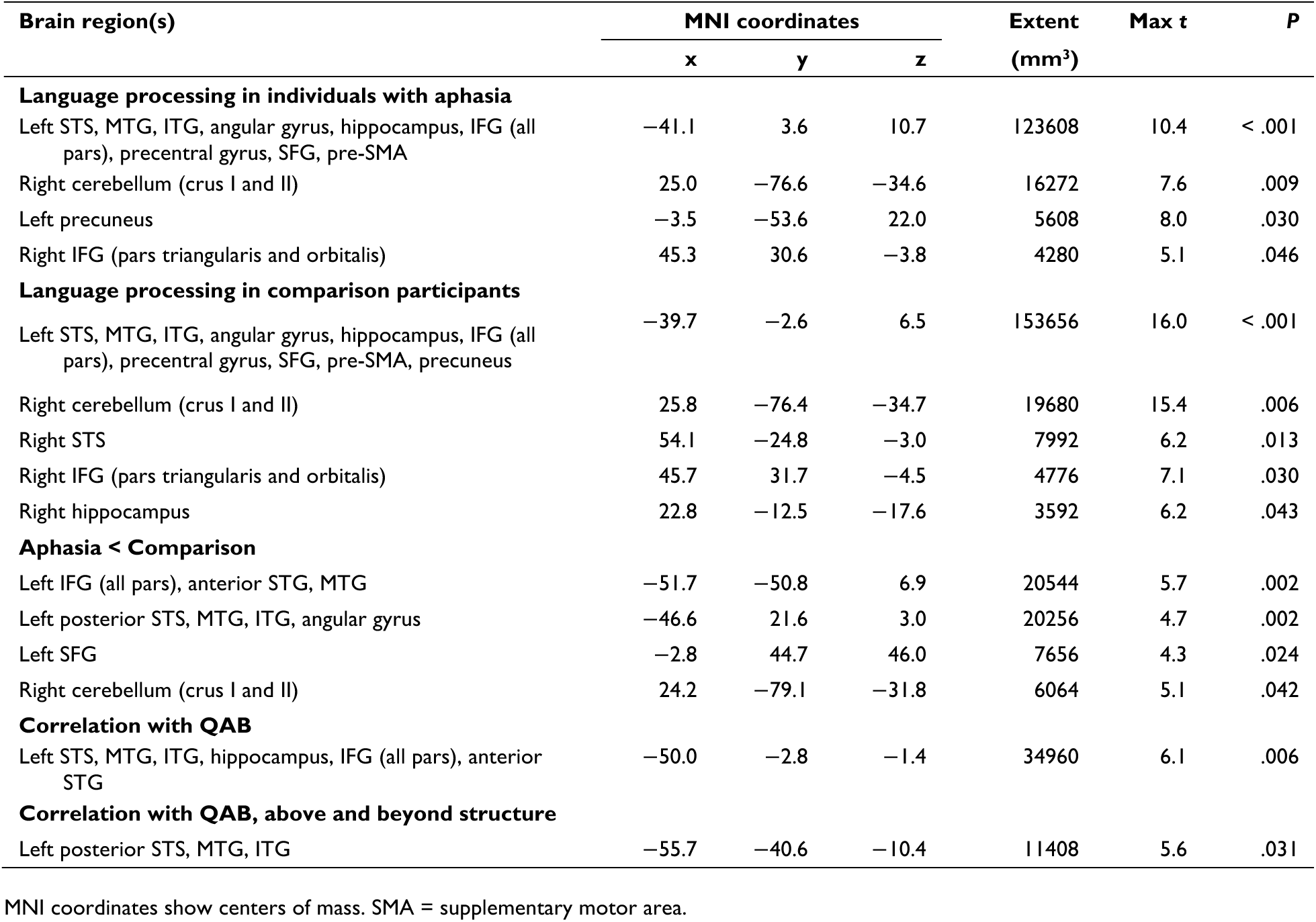
Brain regions activated by each contrast.

Despite the similarity of the activation maps in the two groups, a direct comparison between the groups revealed reduced activation in individuals with aphasia in the left posterior STS extending ventrally to the posterior MTG and posterior ITG, and posteriorly to the angular gyrus; the left IFG (pars orbitalis, triangularis, and opercularis) extending ventrally to the anterior temporal lobe; the medial left superior frontal gyrus; and the right cerebellum (Fig. 3C; Table 3). No regions showed increased activation in individuals with aphasia relative to comparison participants.

The ROI analyses largely mirrored these findings (Supplementary Tables 3−6), revealing reduced activation in individuals with aphasia compared to comparison participants in the left temporal ROI (*β* = −0.25 percent signal change, *t*(109) = −6.23, *P_FDR_* < .001) and left frontal ROI (*β* = −0.22, *t*(109) = −4.04, *P_FDR_* < .001), and no difference between the groups in the right frontal ROI (*β* = −0.008, *t*(109) = −.33, *P_FDR_* = .74). However, the greater sensitivity of the ROI analyses revealed an additional between-groups difference in the right temporal ROI, where individuals with aphasia showed reduced activation relative to comparison participants (*β* = −0.05, *t*(109) = −2.34, *P_FDR_* = .029).

### 3.3 Functional MRI initial correlational analyses

In the initial whole-brain correlational analysis (not yet incorporating structural information), we observed positive associations between voxelwise activity and QAB overall scores in several left-hemisphere language regions, including the left posterior STS extending ventrally to the posterior ITG, and anteriorly to the left anterior STG, and the left IFG (pars orbitalis, triangularis, and opercularis) (Fig. 3D; Table 3). In other words, greater activation in these regions was associated with better aphasia outcomes, operationalized as higher QAB overall scores. No negative relationships were observed.

The ROI analyses largely mirrored these findings (Fig. 3E, Supplementary Tables 7−10), showing positive relationships with QAB overall scores in the left temporal ROI (*β* = 8.80, *t*(62) = 6.26, *P_FDR_* < .001) and left frontal ROI (*β* = 4.56, *t*(62) = 4.23, *P_FDR_* < .001), and no relationship in the right frontal ROI (*β* = 0.16, *t*(62) = 0.05, *P_FDR_* = .96). Again, however, the greater sensitivity of the ROI analyses facilitated the detection of an additional positive relationship between language activation and language function in the right temporal ROI (*β* = 7.92, *t*(62) = 2.13, *P_FDR_* = .049).

By examining the plots demonstrating significant relationships between functional activity and aphasia outcomes in Fig. 3E, it can be readily appreciated that lesion load estimates—depicted with the yellow-orange-red color scale—were strongly associated with both axes of these plots. This indicates that these findings might be driven by structural differences between individuals. To address this potential confound, we next sought to determine whether functional activations have independent explanatory power, or whether their associations with aphasia outcomes are secondary to structural damage to core left hemisphere language regions.

### 3.4 Integration of functional MRI, structural MRI, and behavior

To identify brain regions where activation was associated with successful language processing *above and beyond* what could already be explained by structural damage, we added lesion load (i.e., predicted QAB overall score based on structural imaging alone) to create integrated structure-function-behaviour models. The whole-brain analysis revealed a positive relationship between voxelwise activity and QAB outcomes in a single cluster extending from the left posterior STS to the MTG and ITG (Fig. 4A, Table 3). In this region, greater functional activation was predictive of higher observed QAB overall scores, above and beyond expectations based on structural damage alone. There were no negative associations observed.

**Figure 4.**
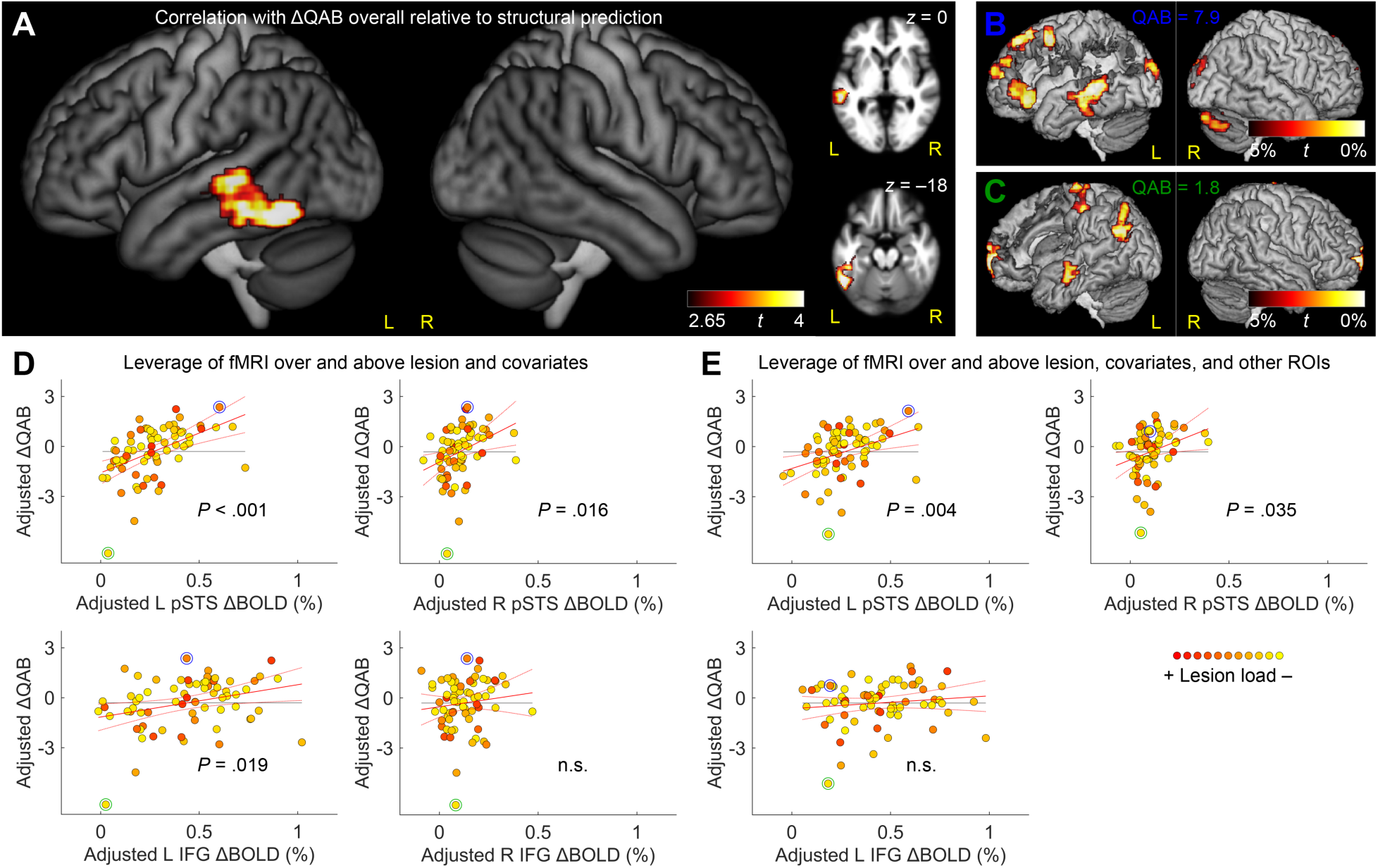
Integrated functional and structural MRI analysis. (**A**) Brain regions where BOLD signal change was correlated with the difference between the actual observed Quick Aphasia Battery (QAB) overall score, and the QAB overall score that was predicted based on lesion location and extent, and covariates. In other words, activation of these regions was predictive of outperforming structural expectations. (**B**) An example of an individual with aphasia who had substantial activation of the left temporal language region, and a QAB score (7.9) that greatly exceeded the score that was predicted based on lesion location and extent. For axial slices and the lesion mask, see Supplementary Fig. 1A. (**C**) An example of an individual with aphasia who had negligible activation of the left temporal language region, and a QAB score (1.8) that was much lower than was predicted based on lesion location and extent. For axial slices and the lesion mask, see Supplementary Fig. 1B. (**D**) Added variable plots (partial regression residual leverage plots) showing the independent predictive value of activation in each of the four regions of interest (ROIs) for predicting QAB overall scores, above and beyond lesion location/extent and other covariates. The color of each participant’s datapoint indicates the lesion load estimate. The case examples from panels C and D are indicated with blue and green circles respectively. n.s. = not significant. (**E**) Added variable plots (partial regression residual leverage plots) showing the extent to which activation in each of the three regions of interest (ROIs) that were predictive in panel D was able to predict QAB overall scores, above and beyond not only structural damage and other covariates, but also the other predictive functional regions.

To illustrate this finding, two case examples are provided (Fig. 4B,C; see Supplementary Fig. 1 for more detail). These individuals both had extensive fronto-parietal lesions reflecting infarction of the upper division of the MCA. Their observed QAB overall scores were quite different (7.9 and 1.8, respectively), which could not be accounted for structurally because of the similarity of their lesions. However, the difference between their aphasia outcomes parallels differences in their functional activation for language processing. The individual in Fig. 4B showed robust and extensive activation in the left posterior temporal lobe, while the individual in Fig. 4C did not, despite this core region being spared in both individuals. These individuals thus exemplify the overall pattern observed in the cohort, whereby activation of the left posterior temporal region was predictive of achieving QAB overall scores above and beyond what was expected based on structural damage.

We next carried out ROI analyses (first, a separate model for each ROI) using the same model structure as in the whole-brain analysis (Fig. 4D, Supplementary Tables 11−14). The ROI analyses confirmed the importance of the left temporal ROI (*β* = 4.87, *t*(60) = 4.39, *P_FDR_* < .001); ancillary analyses confirmed an independent contribution of functional activity in this region when we modeled damage to the left temporal ROI itself instead of the lesion load metric (*β* = 4.59, *t*(60) = 3.72, *P* < .001) or in combination with the lesion load metric (*β* = 4.45, *t*(58) = 3.67, *P* < .001). Furthermore, the greater sensitivity of the ROI analyses revealed additional positive associations between functional activity and aphasia outcomes in the left frontal ROI (*β* = 1.99, *t*(60) = 2.52, *P_FDR_* = .019) (demonstrated in the case examples above) and the right temporal ROI (*β* = 6.18, *t*(60) = 2.74, *P_FDR_* = .016) (not seen in the case examples). There was no association in the right frontal ROI (*β* = 1.72, *t*(60) = 0.88, *P_FDR_* = .38).

While these ROI analyses showed that functional activity in three regions could predict aphasia outcomes above and beyond lesion location and extent, these models were all separate from one another, modeling functional activity in each region in isolation, which is not a realistic assumption. Indeed, when we examined correlations (across participants) between activity in these three regions, we found that in neurologically normal comparison participants, left temporal language activation was strongly correlated with both left frontal language activation (*r* = .56, *P* < .001) and right temporal language activation (*r* = .58, *P* < .001). In the aphasia group, left temporal language activation remained strongly correlated with left frontal language activation (*r* = .58, *P* < .001) and was marginally correlated with right temporal activation (*r* = .22, *P* = .070).

To address this limitation, we carried out a final ROI analysis in which functional activity in the three potentially predictive regions (left temporal, left frontal, right temporal) was entered simultaneously into a single model, along with lesion load (i.e., predicted QAB) and covariates (Fig. 4E; Supplementary Table 15). This model revealed that functional activity in both the left temporal (*β* = 3.75, *t*(58) = 3.00, *P* = .004) and right temporal (*β* = 4.49, *t*(58) = 2.16, *P* = .035) ROIs predicted aphasia outcomes above and beyond all other variables. In contrast, functional activity in the left frontal ROI was no longer predictive (*β* = 0.80, *t*(58) = 1.01, *P* = .32), now that functional activity in the temporal ROIs was included in the same model. To ensure that these findings did not reflect residual confounds of task performance, we repeated the analysis with semantic task accuracy as an additional covariate, and found that the same results held (left temporal *P* = .008; right temporal *P* = .028; left frontal *P* = .82). Similarly, to confirm robustness across lesion quantification methods, we substituted simple lesion extent for our lesion load metric, and the results remained consistent (left temporal *P* < .001; right temporal *P* = .005; left frontal *P* = .47).

The final full model that included functional activity in the left temporal, right temporal, and left frontal regions, lesion load, and covariates, explained 76.8% of the variance in aphasia outcomes (Fig. 5; Supplementary Table 15). In comparison, only 66.5% of the variance was explained when only structural information and covariates were included (Fig. 1C; Supplementary Table 16). In other words, adding distributed functional activity to the model explained an additional 10.3% of the variance in aphasia outcomes (*F*(3, 58) = 8.57, *P* < .001), with a large effect size (Cohen’s *f*^2^ = 0.44).

**Figure 5.**
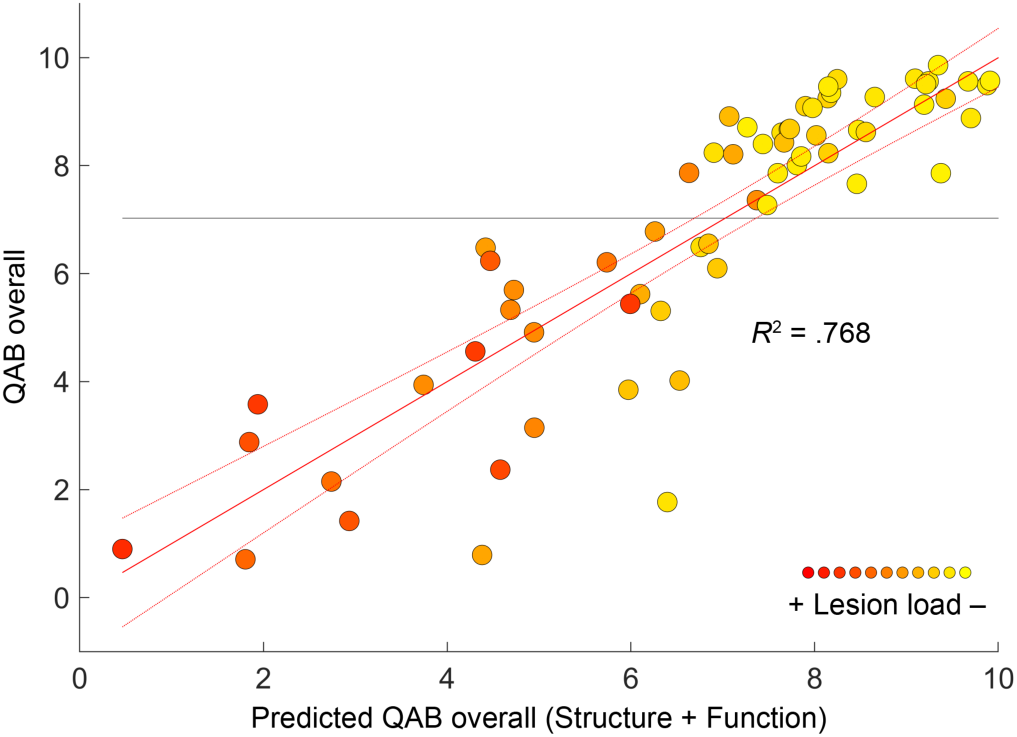
Full model. Prediction of QAB overall from structural data (i.e., lesion load), covariates (age, sex, handedness, education, stroke type, time post onset), and functional activity in the left temporal, right temporal, and left frontal language areas. The color of each participant’s datapoint indicates the lesion load estimate.

Finally, we evaluated the relative contributions of the three functional regions. The left temporal ROI exhibited the largest effect sizes, explaining an additional 8.2% of variance when entered first (Cohen’s *f*^2^ = 0.32, a medium effect), or 3.6% of variance when entered last (*f*^2^ = 0.15, a medium effect). The right temporal ROI explained an additional 3.7% of variance when entered first (*f*^2^ = 0.13, a small effect), or an additional 1.9% when entered last (*f*^2^ = 0.08, a small effect). The left frontal ROI explained an additional 3.2% of variance when entered first (*f*^2^ = 0.11, a small effect), but as noted above, did not make a significant contribution when entered last.

## 4 Discussion

The overall goal of this study was to rigorously and comprehensively characterise the neural basis of language processing in individuals with post-stroke aphasia. We were able to alleviate three major methodological limitations that have often impacted prior studies, and in doing so, we confirmed the two most compelling findings from the literature to date: that individuals with aphasia have reduced activation throughout the left hemisphere language network (Crinion & Price, 2005; Griffis, Nenert, Allendorfer, & Szaflarski, 2017; van Oers et al., 2010) and that greater functional activity throughout the left-hemisphere language network correlates with better aphasia outcomes (Crinion & Price, 2005; Crinion et al., 2006; Fridriksson et al., 2010; Griffis, Nenert, Allendorfer, Vannest, et al., 2017; Papoutsi et al., 2011; Tyler et al., 2011). To build on these findings, we addressed the complex relationships that exist between patterns of structural damage, distributed networks of functional activity in surviving regions, and behaviour, by using machine learning to quantify lesion load, and incorporating this structural information into multivariable analyses of language activations in multiple brain regions and behaviour. These analyses refined our understanding of the roles of left posterior temporal cortex, right posterior temporal cortex, and left inferior frontal cortex in language processing in aphasia, revealing that functional activity in all three of these regions is predictive of aphasia outcomes *above and beyond* patterns of structural damage, and that language activations in both temporal lobes make independent contributions even when functional activity in other regions is taken into account.

### 4.1 Left posterior temporal cortex

Taken together, three of our key findings suggest that left posterior temporal cortex is the most critical region for language processing in post-stroke aphasia. First, in the whole-brain analysis to identify brain regions where language activation predicted aphasia outcomes above and beyond structural information, the only significant cluster was localized to left posterior temporal cortex, extending from the STS to the middle and inferior temporal gyri. This finding was confirmed in our ROI analysis. Second, language activation in the posterior temporal ROI was predictive of aphasia outcomes even when activations in other ROIs (left inferior frontal and right posterior temporal) were included in the same model, demonstrating that the left temporal region makes an independent contribution to aphasia outcomes. Third, left temporal activation explained the greatest proportion of additional variance in aphasia outcomes and always had the largest effect size, regardless of whether it was entered first or last in the multivariable models.

The association of left temporal *damage* with poor aphasia outcomes is well established (Marie, 1906; Metter, 1991; Wilson et al., 2023) and it was not surprising that this effect would be mediated by a reduction in functional activation. But why does functional activation of this region explain language outcomes *above and beyond* structural damage? We propose at least three possible explanations.

First, it may be that our measure of left posterior temporal functional activation provides a more direct measure of the functionality of this region than our lesion load estimate, because our lesion masks may not always reliably encode damage to the critical language region. Reasons for this could include difficulties ascertaining the lesion status of voxels (especially near lesion borders), intersubject variability introduced in the normalization to standard space, and individual differences in the locations of critical language regions.

Second, there may be functional abnormalities in regions that are structurally spared. This could include long term perfusion deficits (Richardson et al., 2011) or diaschitic effects of nearby or distant damage. A structural analysis would not detect these abnormalities and their potentially negative consequences for language outcome, while a functional analysis would be sensitive to reduced or absent activation and would again provide a better estimate of outcome.

The third, and most interesting, possibility is that the functional activation may represent functional reorganization. That is, those individuals who are able to recruit surviving regions within left posterior temporal cortex more extensively than they did before their stroke, or who activate surviving language regions more strongly (resulting in greater BOLD signal change), may have better language outcomes. In a recent longitudinal study, we observed relatively good recoveries for most participants with damage to the left frontal, parietal, and other components of the language network, while individuals with extensive left temporo-parietal damage or complete left perisylvian damage had the poorest outcomes (Wilson et al., 2023). A major mechanism of recovery may involve surviving parts of the language network learning to take over the roles of damaged parts, and the left temporal region may have the greatest capacity in this respect.

The hypothesis that functional reorganization may contribute to our findings can only be verified by future longitudinal studies. To date, there is limited evidence for longitudinal changes over the course of recovery (Wilson & Schneck, 2021). In the most compelling finding so far regarding left temporal cortex, Saur and colleagues reported increased activation in the posterior MTG over the first year after stroke (Saur et al., 2006). In another study from this group, change over time in left MTG activity in a single individual was shown to relate to recovery of language function (Saur & Hartwigsen, 2012). These studies do not distinguish, however, between reactivation of structurally preserved but functionally abnormal tissue versus functional reorganization.

Several previous studies have investigated the relationships between structural damage and functional activity and/or modeled distributed patterns of functional activity (Abel et al., 2015; Fridriksson et al., 2010; Geranmayeh et al., 2016; Griffis et al., 2016; Griffis, Nenert, Allendorfer, & Szaflarski, 2017; Griffis, Nenert, Allendorfer, Vannest, et al., 2017; Johnson et al., 2019; Lorca-Puls et al., 2021; Nenert et al., 2018; Papoutsi et al., 2011; Purcell et al., 2019; Schneider et al., 2022; Sims et al., 2016; Skipper-Kallal et al., 2017a, 2017b; Specht et al., 2009; Stockert et al., 2020; Turkeltaub et al., 2025; Tyler et al., 2010; Warren et al., 2009; Wawrzyniak et al., 2022; Wright et al., 2012). Most notably, in a recent study also using our ALM paradigm, Turkeltaub and colleagues reported that functional activity in several left temporal ROIs (among other regions) was associated with one of the six language measures they investigated—naming and word finding—above and beyond lesion factors (Turkeltaub et al., 2025).

The causality of the functional activations has not been established by our study. Although we have shown that left posterior temporal activation makes a greater contribution to predicting aphasia outcomes than any other region, this does mean that the variable functionality is inherent to this region. The presence or absence of left temporal activity could reflect successful or unsuccessful engagement of the language network, which might ultimately depend on other cognitive, behavioral, or neural factors. For example, an executive deficit (Schumacher et al., 2019) or a pervasive disconnection syndrome (Marebwa et al., 2017; Siegel et al., 2016) could plausibly lead to an inability to recruit the language network for language processing, which could produce the association of functional activity with aphasia outcomes that we observed.

### 4.2 Right posterior temporal cortex

The role of the right temporal lobe in recovery from aphasia is much less well-established. In healthy individuals, the right hemisphere regions homotopic to left hemisphere language areas often show modest activation for language processing (Martin et al., 2022) and contribute to some aspects of language function (Hickok, 2025). This baseline involvement suggests that the right hemisphere is a plausible substrate for reorganization. A seminal study by Weiller et al. (1995) provided the first evidence from functional neuroimaging by demonstrating increased right temporal lobe activity in six individuals who had recovered from Wernicke’s aphasia, relative to comparison participants (albeit with limited statistical support; see Wilson & Schneck, 2021). Later, an important study by Crinion and Price (2005) found that activation of a more anterior right temporal region was associated with language comprehension outcomes in 17 individuals with aphasia. While several other methodologically robust analyses have provided additional support for potential right temporal involvement in language processing after stroke (Leff et al., 2002; Turkeltaub et al., 2025; Tyler et al., 2011), there is a surprising lack of compelling evidence for this position, given the popularity of the claim (Wilson & Schneck, 2021).

Our study provides several pieces of evidence that bear on the role of right posterior temporal cortex in aphasia recovery. First, at the group level, our ROI analyses revealed reduced activation in right posterior temporal cortex in people with aphasia compared to neurotypical participants. This finding is consistent with diaschisis, defined as loss of function in a structurally intact brain region that is connected to damaged regions (Carrera & Tononi, 2014). Two mechanisms could account for this pattern. Most directly, reduced activity could reflect lost excitatory input from damaged left hemisphere homotopic regions, given the robust structural and functional connectivity between left and right temporal cortex (Salvador et al., 2005). We observed this functional connectivity in our comparison participants and, to a lesser extent, in the aphasia group. Alternatively, or additionally, the reduced right hemisphere activation may reflect a more distributed functional consequence: damage to left hemisphere language regions disrupts the entire language network, including the modest right hemisphere contribution to language processing. Diaschisis is an important mechanism in post-stroke aphasia (Price et al., 2001) and has been clearly demonstrated in certain contexts, such as the hyperacute impact of subcortical strokes on cortical language areas (Hillis et al., 2002). However, diaschisis has rarely been convincingly demonstrated in task-based functional neuroimaging studies; exceptions include demonstrations of reduced activity in structurally undamaged left inferior temporal gyrus (Sharp et al., 2004) and anterior temporal regions (Crinion et al., 2006). Notably, our finding of reduced right temporal activity contrasts with most previous reports of increases in right posterior temporal activity in aphasia (Cao et al., 1999; Nenert et al., 2017; Ohyama et al., 1996; Robson et al., 2014; Skipper-Kallal et al., 2017a; Specht et al., 2009; Turkeltaub et al., 2025; Weiller et al., 1995). This discrepancy may reflect task difficulty confounds or other methodological limitations in some prior studies (Wilson & Schneck, 2021), but these factors are unlikely to account for all findings. Instead, the group-level reductions may coexist with compensatory increases in specific individuals or tasks, a pattern that becomes clearer when examining individual variability in our aphasia cohort.

Second, shifting from the group comparison to individual differences, our initial correlational analyses showed that right posterior temporal activation was associated with aphasia outcomes, similar to findings from Crinion and Price (2005), Leff et al. (2002), and Tyler et al. (2011). However, such correlations cannot distinguish whether right temporal activity plays a functional role or simply reflects the status of the left hemisphere: individuals with more severe left hemisphere damage could show both worse outcomes and greater reductions in right temporal activity via diaschisis, thus creating spurious correlations. Turkeltaub et al. (2025) advanced beyond prior work by controlling for left hemisphere lesion factors, showing that right mid-STS activity still predicted one of six language measures—oral word reading—when left hemisphere damage was accounted for. Yet this approach still does not exclude diaschisis mediated through left hemisphere dysfunction rather than structural damage alone.

In this context, our third finding provides stronger evidence for a critical contribution of right posterior temporal cortex to aphasia outcomes after stroke. Specifically, we found that right posterior temporal activation was predictive of aphasia outcomes above and beyond not only lesion load, but even after accounting for functional activation of core left hemisphere language regions. This finding emerged in our analyses based on individually defined functional ROIs, which are more sensitive and better accommodate individual variability than whole-brain analyses. By showing that right temporal activation continued to predict aphasia outcomes even when both lesion load and left hemisphere activation were included in the model, we have demonstrated that the association cannot be explained solely by diaschisis. If right temporal activity were merely a passive reflection of left hemisphere damage and/or dysfunction, it should not provide additional predictive value beyond the variables that encode these factors. Our findings thus support an independent role for right temporal cortex in language processing in aphasia.

We cannot determine based on our data whether the relationship between right temporal activation and aphasia outcomes reflects premorbid variability in the capacity of the non-dominant hemisphere or functional reorganization. Crinion and Price (2005) observed that right temporal activation in the aphasia group in their study did not exceed the range of comparison participants, suggesting that the finding reflected premorbid variability in the capacity of the non-dominant hemisphere rather than reorganization. We also observed that right temporal activation in our aphasia cohort was largely within the range of comparison participants (Fig. 3E), so we, too, consider premorbid variability more likely. However, given that at a group level, people with aphasia had attenuated right temporal activation compared to comparison participants, it is also plausible that our findings reflect functional recruitment of the right hemisphere in a subset of participants, overlaid on a diaschitic effect that reduces right temporal activity at the group level. Future longitudinal studies may be able to resolve this question. In our view, there is presently limited evidence for longitudinal increases in right temporal activity for language (Wilson & Schneck, 2021), but there are some compelling cross-sectional findings (Leff et al., 2002; Turkeltaub et al., 2025) and several recent studies have reported longitudinal increases in right temporal activity for speech (not language) contrasts (Stefaniak et al., 2022; Upton et al., 2024).

### 4.3 Left inferior frontal cortex

The left inferior frontal cortex is the brain region where language activation has most consistently been associated with aphasia outcomes, in several methodologically robust analyses (Fridriksson et al., 2010; Griffis, Nenert, Allendorfer, Vannest, et al., 2017; Tyler et al., 2011) and numerous other studies (Wilson & Schneck, 2021). We replicated this finding (Fig. 3D), and we further observed in our ROI analyses that language activation in the left inferior frontal region predicted aphasia outcomes above and beyond lesion load. This strengthens the evidence for the importance of left inferior frontal activation for language processing in aphasia.

However, surprisingly, when language activations from the bilateral temporal lobes were included in the model, we no longer observed an independent contribution of activity in the left inferior frontal cortex. This may be partly explained by the close anatomical interconnections between left temporal and frontal language regions (Braga et al., 2020; Hampson et al., 2002; Turken & Dronkers, 2011), which led to correlated activations in our data. It is possible that in a larger sample with more representation of different lesion patterns, an independent contribution of frontal activation might be observed. But despite the collinearity, left temporal activations *did* make an independent contribution, above and beyond left frontal activations. This suggests that left temporal activation may be more critical than left frontal activation for language function after stroke.

Why, then, have frontal correlations been more frequently observed than temporal correlations (Wilson & Schneck, 2021)? One possible explanation is that fMRI is inherently biased towards frontal language activations, which are more readily detectable (Diachek et al., 2022) because active tasks, which tend to enhance psychometric properties (Wilson, Yen, et al., 2018) also tend to engage “language executive” regions in the left frontal lobe (Binder et al., 1997).

### 4.4 Other potential mechanisms

Several mechanisms that have been proposed to potentially contribute to language processing in post-stroke aphasia were not strongly supported by our data. Increased activity in the right IFG, particularly in the pars opercularis, is a frequent finding in the literature, including in two methodologically robust studies (Blank et al., 2003; Turkeltaub et al., 2025) and numerous other studies (Wilson & Schneck, 2021). However, we did not observe increased (or decreased) activation in the right IFG in people with aphasia, nor did we find evidence that activity in this region was associated with aphasia outcomes in any of the models that we explored. We suggest that most prior reports of right IFG increases likely reflect recruitment of the multiple demand (MD) network node in the dorsal portion of the pars opercularis, rather than language-specific reorganization in regions homotopic to left hemisphere language areas (Geranmayeh et al., 2014; Quillen et al., 2021). Consistent with this interpretation, Turkeltaub et al. (2025) reported increases specifically in right dorsal IFG consistent with MD network recruitment, though these were not associated with better aphasia outcomes. Our right IFG ROI analysis used a search space that encompassed both language homotopic regions and the MD portion of the dorsal IFG; thus, we cannot determine from this ROI analysis whether there was localized MD recruitment within the right IFG that did not reach statistical significance in our whole-brain analyses.

Beyond the right IFG, we found no evidence of increased MD network activity across the rest of the bilateral MD network in our whole-brain analyses. This is notable because the MD network has been proposed as a potential compensatory mechanism (Brownsett et al., 2014; Geranmayeh et al., 2014, 2017), though our finding aligns with a recent study that also reported no MD upregulation in aphasia (de Clercq et al., 2024).

Finally, we did not observe increased activity in perilesional areas outside the language network (Fridriksson et al., 2012). Although our study did not explicitly investigate perilesional activations with respect to individual lesions, another recent study using our adaptive language mapping paradigm did take this approach, and also did not find evidence for compensatory perilesional activations in aphasia (DeMarco et al., 2022).

### 4.5 Limitations

Our study had several noteworthy limitations. First, the adaptive paradigm did not completely succeed in matching performance across the groups: individuals with aphasia were less accurate on the language task than comparison participants, faced less difficult items, and were slower. Several researchers have similarly made efforts to manipulate and match task difficulty between people with aphasia and comparison participants (Brownsett et al., 2014; Raboyeau et al., 2008; Sharp et al., 2004, 2010) with varying degrees of success. The likely consequences of the language task being more difficult for the individuals with aphasia can be evaluated, since we have examined the impact of linguistic demand in a prior study, and found that linguistic demand modulated the left hemisphere language network, the right IFG, and the bilateral MD network (Quillen et al., 2021). In contrast, here we observed decreased activation in left hemisphere language regions in individuals with aphasia, and no increases in the right IFG, in MD regions, or in any other brain regions. This suggests that the residual performance confounds had a minimal impact and were unlikely to have contributed to our main findings.

Second, although our analyses were multidimensional in many ways, our data reduction choices did leave some variability unexplored in the present study. In particular, we incorporated structural information into our analyses of functional activation and behavior via a single scalar measure of lesion load, which was based only on damage to cortical language regions. It is plausible that lesions to different parts of the language network have different consequences for language processing (Stockert et al., 2020), so future studies should seek to integrate structural information in more specific ways. On the behavioral side, we used a global language measure to characterize language performance, which can be vulnerable to masking important individual variability (Turkeltaub, 2019). However, the measure we used had strong psychometric qualities (Wilson, Eriksson, et al., 2018) and different domains of language processing are highly correlated in aphasia (Hula et al., 2010; Kertesz & Phipps, 1977; Wilson, Eriksson, et al., 2018), so the QAB overall score was a good starting point. Relatedly, our language mapping paradigm and aphasia battery were both focused on core linguistic functions and do not provide information about functional communication abilities in people with aphasia. Future work should investigate the neural correlates of communication in aphasia from a more holistic perspective. Along these lines, although we used the term “aphasia outcomes” throughout this paper, we acknowledge that there are many other important aspects of outcome for people with aphasia beyond purely linguistic function (e.g., functional communication, participation, quality of life).

Third, there were some practical limitations to our cohort of individuals with aphasia. We included participants in the late subacute period (∼3 months post-stroke) despite the potential for continued recovery and neural reorganization throughout the first year (Wilson et al., 2023) and beyond. This decision prioritized adequate sample size while recognizing that language recovery is generally modest after 3 months: in a previous longitudinal study, we observed mean recovery of only 0.4 ± 0.5 QAB points between 3 and 12 months post-stroke (based on 67 patients, including 31 in the present study) (Wilson et al., 2023). We mitigated this limitation by including time post onset in all relevant models, with a logarithmic transformation to match observed recovery trajectories (Yagata et al., 2017). Also, as the data were acquired over a 7-year period, multiple scanners were used with different acquisition parameters. While our preprocessing pipeline was designed to harmonize data across scanners, this is a source of variability that would be better to avoid in future work.

### 4.6 Conclusion

In sum, our goal was to comprehensively characterise the neural basis of language processing in individuals with post-stroke aphasia. We addressed several major methodological challenges by using an adaptive language mapping paradigm with a large and diverse sample of individuals with aphasia, and investigated the complex relationships between patterns of structural damage, distributed networks of functional activity, and behavioral outcomes using multivariable models that integrated structural MRI, functional MRI and behavioral measures. We showed that functional imaging provides critical insights into language processing in aphasia that cannot be obtained from structural imaging alone. Our findings highlighted the importance of the left and right posterior temporal cortices, which we showed make independent contributions to aphasia outcomes after stroke, even after taking into account lesion load and functional activations in other brain regions.

## Data and code availability

The data underlying the analyses reported in this article are available on the Open Science Framework (OSF) at https://osf.io/ga8e5. Specifically, the dataset includes all reported contrast images (thresholded and unthresholded), all individual participant data underlying the ROI and behavioral analyses, and the Matlab code for the statistical analyses.

## Acknowledgements

We gratefully thank the many people with aphasia who generously gave their time and energy to participate in this study; their caregivers and loved ones; Fabi Hersch, Dominique Herrington, Trisha Kennedy, Amanda Hereford, and many other clinicians for assistance with recruitment; Melissa Duff and Catie Chang for helpful feedback; lab members Andrew DeMarco, Sydney Franklin, Ian Quillen, Maysaa Rahman, Jill Rawls, Kiiya Shibata, and Emma Willey for their contributions to this work; and Scott Squire, Seth Smith, and the MRI technologists at the Vanderbilt University Institute of Imaging Science for assistance with MRI scanning.

## Supplementary material

Supplementary material accompanies this article.

