## Supplementary material for "Independent contributions of language activations in left and right temporal cortex to aphasia outcomes after stroke"

**Supplementary Table 1 Demographic and medical history data for all participants**

| PID | Group | Age | Sex | Hand | Educ | Stroke | TPO | Extent |
|---|---|---|---|---|---|---|---|---|
| 0120 | A | 70-74 | M | R | 16 | I | 131.0 | 165.4 |
| 0131 | A | 70-74 | M | R | 18 | I | 116.8 | 134.3 |
| 0182 | A | 70-74 | M | R | 12 | I | 84.3 | 102.4 |
| 0190 | A | 55-59 | M | R | 13 | I | 63.9 | 144.2 |
| 0199 | A | 75-79 | M | R | 16 | I | 62.1 | 86.0 |
| 0308 | A | 50-54 | M | R | 16 | I | 43.1 | 97.7 |
| 0322 | A | 60-64 | M | L | 13 | I | 53.7 | 193.4 |
| 1155 | A | 65-69 | F | R | 13 | H | 103.6 | 11.4 |
| 1295 | A | 30-34 | F | R | 14 | I | 9.5 | 55.5 |
| 1346 | A | 70-74 | M | R | 20 | I | 11.8 | 20.9 |
| 1351 | A | 50-54 | M | R | 12 | I | 12.1 | 10.3 |
| 1354 | A | 65-69 | M | R | 16 | H | 62.5 | 17.5 |
| 1356 | A | 30-34 | F | R | 19 | I | 7.4 | 110.6 |
| 1361 | A | 65-69 | F | R | 15 | I | 82.4 | 53.3 |
| 1368 | A | 70-74 | M | R | 12 | I | 109.6 | 103.9 |
| 1535 | A | 50-54 | M | R | 12 | I | 14.0 | 191.3 |
| 1539 | A | 40-44 | F | R | 18 | I | 12.9 | 190.4 |
| 1554 | A | 45-49 | F | R | 15 | I | 14.2 | 67.3 |
| 1584 | A | 45-49 | F | R | 16 | H | 12.9 | 5.5 |
| 1594 | A | 40-44 | M | R | 14 | I | 12.7 | 29.9 |
| 1619 | A | 50-54 | M | R | 12 | I | 11.8 | 143.9 |
| 1649 | A | 50-54 | M | R | 12 | I | 3.3 | 31.9 |
| 1651 | A | 40-44 | F | R | 16 | I | 3.1 | 65.7 |
| 1656 | A | 80-84 | F | R | 12 | I | 11.9 | 16.3 |
| 1663 | A | 40-44 | F | A | 12 | I | 12.7 | 138.8 |
| 1671 | A | 40-44 | M | R | 12 | H | 12.3 | 17.1 |
| 1686 | A | 55-59 | M | R | 16 | I | 13.1 | 175.8 |
| 1702 | A | 80-84 | M | R | 12 | I | 13.1 | 85.3 |
| 1724 | A | 60-64 | F | R | 14 | I | 3.9 | 11.9 |
| 1731 | A | 45-49 | M | R | 18 | I | 33.1 | 237.2 |
| 1734 | A | 55-59 | M | R | 11 | H | 12.3 | 3.1 |
| 1738 | A | 70-74 | M | R | 14 | I | 101.7 | 164.9 |
| 1742 | A | 55-59 | F | R | 14 | I | 4.3 | 41.1 |
| 1776 | A | 60-64 | M | R | 12 | I | 13.1 | 3.8 |
| 1778 | A | 55-59 | M | R | 20 | I | 9.4 | 19.2 |
| 1812 | A | 75-79 | M | R | 12 | I | 3.2 | 35.4 |
| 1819 | A | 65-69 | F | R | 18 | I | 54.1 | 399.9 |
| 1826 | A | 75-79 | M | R | 12 | I | 2.8 | 2.1 |
| 1847 | A | 55-59 | F | R | 16 | I | 12.0 | 9.3 |
| 1859 | A | 75-79 | F | R | 9 | I | 4.6 | 11.1 |
| 1861 | A | 60-64 | F | R | 13 | I | 12.1 | 0.7 |
| 1894 | A | 60-64 | M | L | 10 | I | 3.2 | 13.9 |
| 1897 | A | 65-69 | M | L | 20 | I | 12.7 | 310.4 |

| | | | | | | | | |
|---|---|---|---|---|---|---|---|---|
| 1915 | A | 50-54 | F | L | 15 | I | 2.9 | 4.9 |
| 1923 | A | 35-39 | F | R | 13 | I | 3.0 | 108.0 |
| 1934 | A | 75-79 | M | R | 12 | I | 3.4 | 7.0 |
| 1936 | A | 35-39 | F | R | 18 | I | 91.9 | 354.1 |
| 1940 | A | 55-59 | F | L | 12 | I | 12.5 | 25.8 |
| 1944 | A | 70-74 | F | R | 16 | I | 143.9 | 78.4 |
| 1970 | A | 75-79 | F | L | 6 | I | 2.9 | 89.9 |
| 1974 | A | 45-49 | M | R | 16 | I | 3.5 | 19.0 |
| 1977 | A | 50-54 | M | R | 10 | I | 3.0 | 15.9 |
| 1980 | A | 55-59 | M | R | 12 | I | 3.1 | 38.7 |
| 1981 | A | 65-69 | F | R | 14 | I | 3.7 | 179.6 |
| 1983 | A | 60-64 | M | R | 16 | I | 14.5 | 103.6 |
| 1996 | A | 40-44 | M | R | 19 | I | 15.6 | 3.0 |
| 2013 | A | 75-79 | M | R | 8 | H | 3.1 | 14.5 |
| 2014 | A | 30-34 | M | R | 12 | H | 3.4 | 18.8 |
| 2018 | A | 80-84 | F | R | 9 | I | 3.3 | 3.5 |
| 2026 | A | 45-49 | M | L | 12 | I | 12.4 | 26.5 |
| 2027 | A | 65-69 | M | R | 16 | I | 18.7 | 38.5 |
| 2039 | A | 55-59 | M | R | 16 | I | 12.1 | 139.1 |
| 2049 | A | 70-74 | F | R | 12 | I | 17.1 | 8.8 |
| 2066 | A | 60-64 | M | R | 16 | I | 17.5 | 17.5 |
| 2105 | A | 45-49 | M | R | 18 | I | 13.5 | 26.2 |
| 2201 | A | 30-34 | F | R | 12 | I | 14.0 | 336.6 |
| 2204 | A | 60-64 | F | R | 12 | I | 3.5 | 78.4 |
| 2220 | A | 60-64 | M | R | 11 | I | 5.0 | 174.5 |
| 2243 | A | 60-64 | M | R | 12 | I | 14.8 | 204.0 |
| 2364 | A | 70-74 | M | A | 20 | I | 259.4 | 434.6 |
| 1087 | C | 40-44 | M | R | 20 | — | — | — |
| 1200 | C | 70-74 | F | R | 16 | — | — | — |
| 1353 | C | 40-44 | M | R | 18 | — | — | — |
| 1362 | C | 35-39 | M | R | 14 | — | — | — |
| 1364 | C | 55-59 | M | L | 18 | — | — | — |
| 1369 | C | 25-29 | M | L | 16 | — | — | — |
| 1370 | C | 80-84 | M | R | 18 | — | — | — |
| 1371 | C | 35-39 | M | R | 18 | — | — | — |
| 1372 | C | 60-64 | F | R | 14 | — | — | — |
| 1373 | C | 55-59 | F | R | 16 | — | — | — |
| 1378 | C | 40-44 | F | L | 18 | — | — | — |
| 1381 | C | 50-54 | F | R | 18 | — | — | — |
| 1387 | C | 65-69 | M | L | 20 | — | — | — |
| 1388 | C | 60-64 | M | R | 20 | — | — | — |
| 1579 | C | 70-74 | M | R | 18 | — | — | — |
| 1580 | C | 55-59 | F | R | 14 | — | — | — |
| 1581 | C | 65-69 | F | L | 16 | — | — | — |
| 1582 | C | 75-79 | F | R | 14 | — | — | — |
| 1583 | C | 70-74 | F | R | 18 | — | — | — |

| | | | | | | | | |
|---|---|---|---|---|---|---|---|---|
| 1587 | C | 75-79 | F | L | 20 | — | — | — |
| 1595 | C | 55-59 | M | R | 16 | — | — | — |
| 1607 | C | 35-39 | M | R | 12 | — | — | — |
| 1608 | C | 20-24 | F | R | 18 | — | — | — |
| 1621 | C | 45-49 | M | R | 18 | — | — | — |
| 1623 | C | 45-49 | F | A | 18 | — | — | — |
| 1624 | C | 60-64 | F | R | 16 | — | — | — |
| 1626 | C | 60-64 | M | L | 16 | — | — | — |
| 1627 | C | 55-59 | F | R | 16 | — | — | — |
| 1628 | C | 60-64 | F | R | 16 | — | — | — |
| 1740 | C | 60-64 | M | R | 20 | — | — | — |
| 1798 | C | 20-24 | M | L | 16 | — | — | — |
| 1811 | C | 50-54 | F | R | 18 | — | — | — |
| 1818 | C | 60-64 | F | R | 16 | — | — | — |
| 1827 | C | 60-64 | F | R | 14 | — | — | — |
| 1835 | C | 40-44 | M | R | 18 | — | — | — |
| 1839 | C | 40-44 | M | R | 16 | — | — | — |
| 1841 | C | 40-44 | F | R | 16 | — | — | — |
| 1842 | C | 40-44 | F | R | 16 | — | — | — |
| 1844 | C | 80-84 | F | L | 20 | — | — | — |
| 1849 | C | 60-64 | F | R | 16 | — | — | — |
| 1850 | C | 50-54 | M | R | 16 | — | — | — |
| 1851 | C | 40-44 | F | R | 16 | — | — | — |
| 1852 | C | 80-84 | F | A | 16 | — | — | — |
| 1873 | C | 20-24 | F | R | 17 | — | — | — |
| 1905 | C | 70-74 | M | R | 16 | — | — | — |

PID = Participant identifier; Group: A = Aphasia; C = Comparison; Sex: M = Male; F = Female; Hand = Handedness: R = Right; L = Left; A = Ambidextrous; Educ = Education (years); Stroke: I = Ischemic; H = Hemorrhagic; TPO = Time post onset (months); Extent = Lesion extent (cubic centimeters).

**Supplementary Table 2 Quick Aphasia Battery subscores for all participants**

| PID | Group | WC | SC | WF | GC | SMP | Rep | Read | QAB | WWC | PPT |
|---|---|---|---|---|---|---|---|---|---|---|---|
| 0120 | A | 9.86 | 3.47 | 4.17 | 6.96 | 8.33 | 6.53 | 5.14 | 6.24 | 9.58 | 14 |
| 0131 | A | 10 | 1.53 | 1.25 | 6.50 | 7.50 | 3.61 | 3.61 | 4.91 | 10 | 14 |
| 0182 | A | 9.58 | 5.28 | 2.50 | 0.38 | 0.83 | 4.44 | 3.20 | 4.02 | 10 | 14 |
| 0190 | A | 10 | 6.81 | 6.81 | 7.88 | 9.17 | 7.64 | 8.89 | 7.87 | 10 | 13 |
| 0199 | A | 10 | 7.09 | 4.58 | 3.46 | 0.00 | 4.17 | 4.45 | 5.31 | 10 | 14 |
| 0308 | A | 9.31 | 3.06 | 3.75 | 6.63 | 10 | 3.33 | 4.03 | 5.70 | 10 | 14 |
| 0322 | A | 5.42 | 0.83 | 0.17 | 0.04 | 0.00 | 3.34 | 0.00 | 1.42 | 5.83 | 13 |
| 1155 | A | 10 | 9.86 | 8.83 | 9.38 | 10 | 9.86 | 9.72 | 9.46 | 10 | 14 |
| 1295 | A | 9.86 | 7.78 | 7.94 | 5.42 | 8.33 | 6.25 | 6.11 | 7.41 | 10 | 14 |
| 1346 | A | 10 | 8.20 | 7.25 | 8.54 | 10 | 8.75 | 9.58 | 8.62 | 10 | 14 |
| 1351 | A | 8.89 | 2.22 | 6.64 | 7.96 | 10 | 4.72 | 6.80 | 6.49 | 10 | 14 |
| 1354 | A | 10 | 6.39 | 4.92 | 8.88 | 10 | 7.64 | 8.61 | 7.66 | 10 | 14 |
| 1356 | A | 9.30 | 1.39 | 2.42 | 0.79 | 0.00 | 2.78 | 1.80 | 3.15 | 10 | 14 |
| 1361 | A | 9.58 | 3.06 | 4.78 | 7.80 | 10 | 5.83 | 6.11 | 6.55 | 10 | 14 |
| 1368 | A | 9.44 | 6.81 | 7.78 | 8.55 | 10 | 7.64 | 9.86 | 8.23 | 10 | 14 |
| 1535 | A | 9.79 | 1.25 | 2.75 | 0.13 | 2.50 | 4.58 | 2.50 | 3.58 | 10 | 14 |
| 1539 | A | 9.17 | 7.92 | 6.50 | 7.38 | 5.00 | 7.92 | 7.92 | 7.36 | 8.75 | 14 |
| 1554 | A | 10 | 9.79 | 7.50 | 5.00 | 7.50 | 8.33 | 7.50 | 7.86 | 10 | 14 |
| 1584 | A | 10 | 10 | 7.50 | 9.63 | 10 | 9.17 | 9.58 | 9.29 | 10 | 14 |
| 1594 | A | 10 | 9.17 | 9.75 | 10 | 10 | 10 | 10 | 9.61 | 10 | 14 |
| 1619 | A | 8.96 | 0.21 | 3.00 | 5.00 | 2.50 | 4.17 | 0.83 | 3.94 | 7.08 | 14 |
| 1649 | A | 10 | 10 | 9.00 | 9.25 | 10 | 8.33 | 7.92 | 9.25 | — | — |
| 1651 | A | 10 | 10 | 7.00 | 9.25 | 10 | 9.17 | 9.58 | 8.91 | — | — |
| 1656 | A | 10 | 7.50 | 9.00 | 9.63 | 7.50 | 8.75 | 8.33 | 8.66 | 9.58 | 14 |
| 1663 | A | 9.79 | 10 | 8.75 | 9.75 | 10 | 9.17 | 7.08 | 9.24 | 10 | 14 |
| 1671 | A | 8.75 | 4.17 | 9.00 | 9.63 | 10 | 8.33 | 8.33 | 8.01 | 10 | 14 |
| 1686 | A | 8.33 | 2.50 | 0.00 | 0.00 | 0.00 | 2.50 | 0.00 | 2.15 | 5.83 | 14 |
| 1702 | A | 9.79 | 9.58 | 6.50 | 8.88 | 10 | 9.17 | 7.50 | 8.71 | 10 | 12 |
| 1724 | A | 10 | 10 | 9.50 | 9.38 | 7.50 | 8.75 | 9.17 | 9.27 | 10 | 14 |
| 1731 | A | 9.58 | 1.67 | 3.00 | 0.75 | 7.50 | 5.42 | 2.92 | 4.34 | 10 | 13 |
| 1734 | A | 9.38 | 7.71 | 7.75 | 9.63 | 10 | 7.50 | 7.50 | 8.40 | 7.50 | 14 |
| 1738 | A | 10 | 8.33 | 8.50 | 8.75 | 5.00 | 7.92 | 9.17 | 8.21 | 10 | 14 |
| 1742 | A | 9.17 | 6.04 | 8.50 | 9.50 | 10 | 8.75 | 9.58 | 8.56 | 10 | 14 |
| 1776 | A | 10 | 7.92 | 8.25 | 9.00 | 10 | 8.33 | 3.75 | 8.61 | 8.75 | 14 |
| 1778 | A | 10 | 10 | 7.75 | 8.25 | 10 | 9.17 | 8.33 | 8.88 | 10 | 14 |
| 1812 | A | 10 | 3.33 | 2.50 | 7.50 | 10 | 7.50 | 5.42 | 6.10 | 6.25 | 13 |
| 1819 | A | 4.58 | 0.00 | 0.00 | 0.63 | 7.50 | 5.00 | 0.42 | 2.37 | 1.67 | 11 |
| 1826 | A | 10 | 7.71 | 5.08 | 8.38 | 10 | 9.58 | 10 | 8.17 | 8.33 | 13 |
| 1847 | A | 9.79 | 8.75 | 8.75 | 9.63 | 10 | 9.17 | 9.58 | 9.10 | 10 | 14 |
| 1859 | A | 9.79 | 6.09 | 9.75 | 9.38 | 10 | 8.75 | 5.42 | 8.67 | 7.08 | 14 |
| 1861 | A | 10 | 10 | 9.33 | 9.50 | 10 | 9.17 | 9.17 | 9.49 | 10 | 14 |
| 1894 | A | 9.79 | 10 | 10 | 9.25 | 10 | 8.33 | 7.92 | 9.56 | 10 | 14 |
| 1897 | A | 4.38 | 0.63 | 0.00 | 0.00 | 0.00 | 0.00 | 0.00 | 0.90 | 0.00 | 11 |

| | | | | | | | | | | |
|---|---|---|---|---|---|---|---|---|---|---|
| 1915 | A | 10 | 8.96 | 9.25 | 9.25 | 10 | 9.17 | 8.33 | 9.35 | 10 | 14 |
| 1923 | A | 8.33 | 3.33 | 2.75 | 6.50 | 10 | 5.00 | 0.83 | 5.33 | 9.58 | 14 |
| 1934 | A | 9.17 | 7.71 | 5.75 | 8.38 | 10 | 8.75 | 7.08 | 7.86 | 0.83 | 13 |
| 1936 | A | 9.79 | 4.38 | 5.25 | 4.63 | 10 | 4.17 | 5.83 | 6.21 | 7.92 | 13 |
| 1940 | A | 10 | 10 | 9.83 | 9.25 | 10 | 9.17 | 10 | 9.60 | 10 | 14 |
| 1944 | A | 10 | 8.54 | 7.83 | 8.25 | 7.50 | 9.17 | 9.17 | 8.24 | 10 | 14 |
| 1970 | A | 4.38 | 0.63 | 2.33 | 5.00 | 10 | 2.92 | — | 3.85 | 2.50 | 11 |
| 1974 | A | 9.79 | 9.58 | 10 | 9.25 | 10 | 8.33 | 9.00 | 9.57 | 10 | 14 |
| 1977 | A | 9.38 | 2.29 | 8.00 | 8.25 | 10 | 7.50 | 7.50 | 7.27 | 8.75 | 13 |
| 1980 | A | 10 | 4.17 | 4.42 | 7.00 | 10 | 5.00 | 7.50 | 6.48 | 8.33 | 14 |
| 1981 | A | 3.75 | 0.00 | 0.00 | 0.00 | 0.00 | 0.00 | 0.42 | 0.71 | 6.25 | 10 |
| 1983 | A | 10 | 8.96 | 8.08 | 8.50 | 7.50 | 8.33 | 8.75 | 8.43 | 10 | 14 |
| 1996 | A | 10 | 10 | 10 | 9.75 | 10 | 10 | 9.17 | 9.90 | 10 | 14 |
| 2013 | A | 9.79 | 9.17 | 8.08 | 9.50 | 10 | 9.17 | 9.17 | 9.13 | 10 | 14 |
| 2014 | A | 10 | 10 | 10 | 9.63 | 10 | 9.58 | 7.50 | 9.66 | 10 | 14 |
| 2018 | A | 10 | 9.17 | 9.75 | 9.38 | 10 | 8.75 | 9.17 | 9.56 | 9.58 | 14 |
| 2026 | A | 9.79 | 8.75 | 9.00 | 9.50 | 10 | 10 | 7.50 | 9.07 | 10 | 14 |
| 2027 | A | 10 | 10 | 10 | 9.75 | 10 | 10 | 9.17 | 9.90 | 9.58 | 14 |
| 2039 | A | 9.58 | 3.75 | 1.50 | 6.63 | 10 | 4.58 | 3.75 | 5.62 | 7.92 | 13 |
| 2049 | A | 10 | 9.79 | 9.00 | 9.75 | 10 | 9.17 | 10 | 9.51 | 10 | 14 |
| 2066 | A | 10 | 7.71 | 8.50 | 8.75 | 10 | 8.33 | 8.33 | 8.68 | 10 | 14 |
| 2105 | A | 10 | 9.79 | 10 | 9.75 | 10 | 10 | 9.17 | 9.86 | 10 | 14 |
| 2201 | A | 8.75 | 0.83 | 1.50 | 0.00 | 5.00 | 4.17 | 0.00 | 2.88 | 8.75 | 12 |
| 2204 | A | 7.92 | 4.79 | 7.00 | 7.13 | 10 | 5.00 | 7.92 | 6.78 | 10 | 14 |
| 2220 | A | 2.92 | 1.46 | 0.00 | 0.00 | 0.00 | 0.00 | 0.00 | 0.79 | 0.00 | 11 |
| 2243 | A | 6.88 | 1.04 | 1.00 | 0.00 | 0.00 | 2.50 | 0.00 | 1.77 | 2.92 | 11 |
| 2364 | A | 9.79 | 0.00 | 5.25 | 4.13 | 7.50 | 7.08 | 7.08 | 5.44 | 9.58 | 14 |
| 1087 | C | 10 | 10 | 10 | 10 | 10 | 10 | 10 | 10 | — | — |
| 1200 | C | 10 | 9.58 | 10 | 9.63 | 10 | 9.58 | 9.17 | 9.77 | — | — |
| 1353 | C | 10 | 10 | 10 | 10 | 10 | 10 | 10 | 10 | — | — |
| 1362 | C | 10 | 5.42 | 10 | 9.50 | 10 | 9.17 | 9.17 | 8.97 | — | — |
| 1364 | C | 10 | 10 | 9.00 | 10 | 10 | 10 | 10 | 9.86 | — | — |
| 1369 | C | 10 | 9.17 | 10 | 9.63 | 10 | 9.17 | 9.58 | 9.65 | — | — |
| 1370 | C | 10 | 9.58 | 10 | 10 | 10 | 10 | 10 | 9.92 | — | — |
| 1371 | C | 10 | 10 | 10 | 9.88 | 10 | 10 | 9.58 | 9.95 | — | — |
| 1372 | C | 10 | 9.17 | 9.25 | 10 | 10 | 10 | 10 | 9.74 | — | — |
| 1373 | C | 10 | 7.50 | 9.75 | 9.63 | 10 | 8.75 | 9.17 | 9.30 | — | — |
| 1378 | C | 10 | 10 | 8.50 | 9.50 | 10 | 10 | 10 | 9.72 | — | — |
| 1381 | C | 10 | 10 | 10 | 10 | 10 | 10 | 8.75 | 9.90 | — | — |
| 1387 | C | 10 | 10 | 10 | 9.75 | 10 | 9.17 | 10 | 9.90 | — | — |
| 1388 | C | 10 | 10 | 10 | 10 | 10 | 10 | 10 | 10 | — | — |
| 1579 | C | 10 | 10 | 10 | 10 | 10 | 10 | 10 | 10 | — | — |
| 1580 | C | 10 | 10 | 10 | 10 | 10 | 10 | 10 | 10 | — | — |
| 1581 | C | 10 | 10 | 10 | 10 | 10 | 10 | 10 | 10 | — | — |
| 1582 | C | 10 | 10 | 10 | 10 | 10 | 10 | 10 | 10 | — | — |
| 1583 | C | 10 | 10 | 10 | 10 | 10 | 10 | 10 | 10 | — | — |

| | | | | | | | | | | | |
|---|---|---|---|---|---|---|---|---|---|---|---|
| 1587 | C | 9.58 | 10 | 10 | 9.75 | 10 | 10 | 9.17 | 9.82 | — | — |
| 1595 | C | 8.75 | 10 | 10 | 9.75 | 10 | 9.17 | 10 | 9.67 | — | — |
| 1607 | C | 10 | 10 | 9.75 | 9.88 | 10 | 8.75 | 10 | 9.85 | — | — |
| 1608 | C | 10 | 10 | 10 | 10 | 10 | 10 | 10 | 10 | — | — |
| 1621 | C | 10 | 8.33 | 10 | 10 | 10 | 10 | 10 | 9.70 | — | — |
| 1623 | C | 10 | 9.58 | 10 | 10 | 10 | 10 | 10 | 9.92 | — | — |
| 1624 | C | 10 | 10 | 10 | 10 | 10 | 10 | 10 | 10 | — | — |
| 1626 | C | 10 | 6.67 | 9.75 | 9.88 | 10 | 9.58 | 10 | 9.31 | — | — |
| 1627 | C | 10 | 10 | 10 | 10 | 10 | 10 | 10 | 10 | — | — |
| 1628 | C | 10 | 10 | 10 | 10 | 10 | 10 | 10 | 10 | — | — |
| 1740 | C | 10 | 10 | 10 | 10 | 10 | 10 | 10 | 10 | — | — |
| 1798 | C | 10 | 10 | 10 | 10 | 10 | 10 | 10 | 9.79 | — | — |
| 1811 | C | 10 | 10 | 10 | 10 | 10 | 10 | 10 | 10 | — | — |
| 1818 | C | 10 | 8.75 | 9.00 | 10 | 10 | 10 | 10 | 9.64 | — | — |
| 1827 | C | 10 | 10 | 10 | 10 | 10 | 10 | 10 | 10 | — | — |
| 1835 | C | 10 | 10 | 10 | 10 | 10 | 10 | 10 | 10 | — | — |
| 1839 | C | 10 | 9.58 | 10 | 10 | 10 | 10 | 10 | 9.92 | — | — |
| 1841 | C | 10 | 10 | 10 | 10 | 10 | 10 | 10 | 10 | — | — |
| 1842 | C | 9.58 | 10 | 9.00 | 10 | 10 | 10 | 10 | 9.79 | — | — |
| 1844 | C | 9.58 | 8.75 | 10 | 10 | 10 | 10 | 10 | 9.70 | — | — |
| 1849 | C | 10 | 10 | 10 | 10 | 10 | 10 | 10 | 10 | — | — |
| 1850 | C | 10 | 9.17 | 10 | 10 | 10 | 10 | 10 | 9.85 | — | — |
| 1851 | C | 10 | 10 | 9.25 | 10 | 10 | 10 | 10 | 9.90 | — | — |
| 1852 | C | 10 | 8.75 | 10 | 10 | 10 | 10 | 10 | 9.78 | — | — |
| 1873 | C | 10 | 10 | 10 | 10 | 10 | 10 | 10 | 10 | — | — |
| 1905 | C | 10 | 9.58 | 10 | 10 | 10 | 10 | 10 | 9.92 | — | — |

PID = Participant identifier; Group: A = Aphasia; C = Comparison; WC = Word comprehension; SC = Sentence comprehension; WF = Word finding; GC = Grammatical construction; SMP = Speech motor programming; Rep = Repetition; Read = Reading aloud; QAB = Quick Aphasia Battery overall; WWC = Written word comprehension; PPT = Pyramids and Palm Trees—Pictures.

**Supplementary Table 3  Between-groups comparison of left temporal activation**

| | Estimate | SE | *t* | *P* |
|---|---|---|---|---|
| (Intercept) | 0.75711 | 0.13864 | 5.4609 | 3.0108e-07 |
| Aphasia | -0.25102 | 0.040292 | -6.23 | 8.9669e-09 |
| Age | -0.0017091 | 0.0011818 | -1.4462 | 0.151 |
| SexF | -0.025318 | 0.034688 | -0.72988 | 0.46703 |
| HandLA | 0.0071371 | 0.045326 | 0.15746 | 0.87517 |
| Educ | -0.0091274 | 0.0065136 | -1.4013 | 0.16397 |

Linear regression model: L_Temporal ~ 1 + Aphasia + Age + SexF + HandLA + Educ
Number of observations: 115, Error degrees of freedom: 109
Root Mean Squared Error: 0.182
R-squared: 0.306,  Adjusted R-Squared: 0.274
F-statistic vs. constant model: 9.6, p-value = 1.3e-07

**Supplementary Table 4  Between-groups comparison of left frontal activation**

| | Estimate | SE | *t* | *P* |
|---|---|---|---|---|
| (Intercept) | 1.0928 | 0.18497 | 5.9083 | 4.004e-08 |
| Aphasia | -0.21719 | 0.053755 | -4.0405 | 9.963e-05 |
| Age | -0.0030667 | 0.0015767 | -1.9451 | 0.054342 |
| SexF | 0.066307 | 0.046279 | 1.4328 | 0.15478 |
| HandLA | -0.068167 | 0.060471 | -1.1273 | 0.26211 |
| Educ | -0.019403 | 0.00869 | -2.2328 | 0.027607 |

Linear regression model: L_Frontal ~ 1 + Aphasia + Age + SexF + HandLA + Educ
Number of observations: 115, Error degrees of freedom: 109
Root Mean Squared Error: 0.243
R-squared: 0.201,  Adjusted R-Squared: 0.164
F-statistic vs. constant model: 5.47, p-value = 0.000157

**Supplementary Table 5  Between-groups comparison of right temporal activation**

| | Estimate | SE | *t* | *P* |
|---|---|---|---|---|
| (Intercept) | 0.15946 | 0.072479 | 2.2001 | 0.029909 |
| Aphasia | -0.049196 | 0.021064 | -2.3356 | 0.021344 |
| Age | -0.0006178 | 0.00061781 | -0.99998 | 0.31954 |
| SexF | 0.00042648 | 0.018134 | 0.023518 | 0.98128 |
| HandLA | 0.0027412 | 0.023696 | 0.11568 | 0.90812 |
| Educ | 0.002065 | 0.0034052 | 0.60644 | 0.54548 |

Linear regression model: R_Temporal ~ 1 + Aphasia + Age + SexF + HandLA + Educ
Number of observations: 115, Error degrees of freedom: 109
Root Mean Squared Error: 0.095
R-squared: 0.0981,  Adjusted R-Squared: 0.0568
F-statistic vs. constant model: 2.37, p-value = 0.0438

**Supplementary Table 6  Between-groups comparison of right frontal activation**

| | Estimate | SE | *t* | *P* |
|---|---|---|---|---|
| (Intercept) | 0.1855 | 0.08394 | 2.2099 | 0.029199 |
| Aphasia | -0.0080375 | 0.024395 | -0.32948 | 0.74243 |
| Age | -0.0013727 | 0.00071551 | -1.9184 | 0.057675 |
| SexF | 0.065028 | 0.021002 | 3.0963 | 0.0024916 |
| HandLA | 0.045105 | 0.027443 | 1.6436 | 0.10314 |
| Educ | -0.00014274 | 0.0039437 | -0.036194 | 0.97119 |

Linear regression model: R_Frontal ~ 1 + Aphasia + Age + SexF + HandLA + Educ
Number of observations: 115, Error degrees of freedom: 109
Root Mean Squared Error: 0.11
R-squared: 0.141,  Adjusted R-Squared: 0.102
F-statistic vs. constant model: 3.58, p-value = 0.00491

**Supplementary Table 7 Initial correlation of left temporal activation with aphasia outcome**

| | Estimate | SE | *t* | *P* |
|---|---|---|---|---|
| (Intercept) | 2.7403 | 2.1316 | 1.2855 | 0.20338 |
| Age | 0.015457 | 0.020707 | 0.74645 | 0.45822 |
| SexF | 0.30123 | 0.52295 | 0.57602 | 0.56669 |
| HandLA | -0.159 | 0.7547 | -0.21067 | 0.83383 |
| Educ | 0.099781 | 0.10058 | 0.99204 | 0.32504 |
| StrHem | 2.3536 | 0.86895 | 2.7085 | 0.0087241 |
| LogTPO | -0.27629 | 0.24194 | -1.142 | 0.25785 |
| L_Temporal | 8.7974 | 1.4048 | 6.2625 | 3.9881e-08 |

Linear regression model: QAB ~ 1 + Age + SexF + HandLA + Educ + StrHem + LogTPO + L_Temporal
Number of observations: 70, Error degrees of freedom: 62
Root Mean Squared Error: 2.08
R-squared: 0.443, Adjusted R-Squared: 0.38
F-statistic vs. constant model: 7.03, p-value = 3.6e-06

**Supplementary Table 8 Initial correlation of left frontal activation with aphasia outcome**

| | Estimate | SE | *t* | *P* |
|---|---|---|---|---|
| (Intercept) | 3.0758 | 2.4404 | 1.2604 | 0.21226 |
| Age | 0.020364 | 0.023382 | 0.87089 | 0.38717 |
| SexF | -0.050523 | 0.60208 | -0.083915 | 0.93339 |
| HandLA | -0.057972 | 0.85242 | -0.068009 | 0.946 |
| Educ | 0.097119 | 0.11361 | 0.85482 | 0.39594 |
| StrHem | 2.1425 | 0.97715 | 2.1926 | 0.032099 |
| LogTPO | -0.30933 | 0.27222 | -1.1363 | 0.26019 |
| L_Frontal | 4.5557 | 1.0774 | 4.2284 | 7.8838e-05 |

Linear regression model: QAB ~ 1 + Age + SexF + HandLA + Educ + StrHem + LogTPO + L_Frontal
Number of observations: 70, Error degrees of freedom: 62
Root Mean Squared Error: 2.34
R-squared: 0.294, Adjusted R-Squared: 0.214
F-statistic vs. constant model: 3.68, p-value = 0.00218

**Supplementary Table 9 Initial correlation of right temporal activation with aphasia outcome**

| | Estimate | SE | *t* | *P* |
|---|---|---|---|---|
| (Intercept) | 6.3893 | 2.5235 | 2.5319 | 0.013892 |
| Age | 0.011253 | 0.025534 | 0.44069 | 0.66097 |
| SexF | 0.24086 | 0.65558 | 0.3674 | 0.71457 |
| HandLA | -0.49972 | 0.93041 | -0.5371 | 0.59312 |
| Educ | -0.0085561 | 0.12479 | -0.068564 | 0.94556 |
| StrHem | 1.6035 | 1.0956 | 1.4636 | 0.14836 |
| LogTPO | -0.35259 | 0.29796 | -1.1833 | 0.24119 |
| R_Temporal | 7.9269 | 3.7177 | 2.1322 | 0.036956 |

Linear regression model: QAB ~ 1 + Age + SexF + HandLA + Educ + StrHem + LogTPO + R_Temporal
Number of observations: 70, Error degrees of freedom: 62
Root Mean Squared Error: 2.56
R-squared: 0.152, Adjusted R-Squared: 0.0564
F-statistic vs. constant model: 1.59, p-value = 0.155

**Supplementary Table 10 Initial correlation of right frontal activation with aphasia outcome**

| | Estimate | SE | *t* | *P* |
|---|---|---|---|---|
| (Intercept) | 6.4634 | 2.6643 | 2.4259 | 0.018192 |
| Age | 0.012489 | 0.027086 | 0.46107 | 0.64636 |
| SexF | 0.49067 | 0.72767 | 0.67431 | 0.50262 |
| HandLA | -0.41988 | 0.98145 | -0.42782 | 0.67027 |
| Educ | 0.032141 | 0.12778 | 0.25153 | 0.80224 |
| StrHem | 2.0985 | 1.1096 | 1.8912 | 0.063271 |
| LogTPO | -0.3823 | 0.31134 | -1.2279 | 0.22412 |
| R_Frontal | 0.16365 | 3.1403 | 0.052113 | 0.95861 |

Linear regression model: QAB ~ 1 + Age + SexF + HandLA + Educ + StrHem + LogTPO + R_Frontal
Number of observations: 70, Error degrees of freedom: 62
Root Mean Squared Error: 2.65
R-squared: 0.09, Adjusted R-Squared: -0.0127
F-statistic vs. constant model: 0.876, p-value = 0.53

**Supplementary Table 11 Contribution of left temporal activation to aphasia outcome, above and beyond structural damage**

| | Estimate | SE | *t* | *P* |
|---|---|---|---|---|
| (Intercept) | -7.1316 | 2.094 | -3.4058 | 0.0011824 |
| Age | -0.015389 | 0.014667 | -1.0492 | 0.29831 |
| SexF | 0.5928 | 0.36614 | 1.619 | 0.11069 |
| HandLA | -0.27405 | 0.54504 | -0.5028 | 0.61695 |
| Educ | 0.1194 | 0.069406 | 1.7204 | 0.09052 |
| StrHem | 0.69253 | 0.63643 | 1.0881 | 0.28088 |
| LogTPO | 2.0451 | 0.52541 | 3.8923 | 0.0002518 |
| PredQAB | 1.5589 | 0.22767 | 6.8471 | 4.5586e-09 |
| LogTPOxPredQAB | -0.27089 | 0.069664 | -3.8885 | 0.00025498 |
| L_Temporal | 4.8671 | 1.1087 | 4.39 | 4.6713e-05 |

Linear regression model: QAB ~ 1 + Age + SexF + HandLA + Educ + StrHem + LogTPO + PredQAB + LogTPOxPredQAB + L_Temporal
Number of observations: 70, Error degrees of freedom: 60
Root Mean Squared Error: 1.42
R-squared: 0.746, Adjusted R-Squared: 0.708
F-statistic vs. constant model: 19.6, p-value = 8.44e-15

**Supplementary Table 12 Contribution of left frontal activation to aphasia outcome, above and beyond structural damage**

| | Estimate | SE | *t* | *P* |
|---|---|---|---|---|
| (Intercept) | -7.3492 | 2.3033 | -3.1907 | 0.0022582 |
| Age | -0.016852 | 0.016124 | -1.0451 | 0.30016 |
| SexF | 0.44514 | 0.40934 | 1.0874 | 0.28119 |
| HandLA | -0.16017 | 0.59579 | -0.26883 | 0.78898 |
| Educ | 0.12096 | 0.076071 | 1.5901 | 0.11706 |
| StrHem | 0.348 | 0.68682 | 0.50668 | 0.61424 |
| LogTPO | 2.0459 | 0.57428 | 3.5625 | 0.00072698 |
| PredQAB | 1.6539 | 0.24711 | 6.6929 | 8.3365e-09 |
| LogTPOxPredQAB | -0.26585 | 0.076134 | -3.4918 | 0.00090665 |
| L_Frontal | 1.9893 | 0.78869 | 2.5222 | 0.014332 |

Linear regression model: QAB ~ 1 + Age + SexF + HandLA + Educ + StrHem + LogTPO + PredQAB + LogTPOxPredQAB + L_Frontal
Number of observations: 70, Error degrees of freedom: 60
Root Mean Squared Error: 1.56
R-squared: 0.697, Adjusted R-Squared: 0.651
F-statistic vs. constant model: 15.3, p-value = 1.39e-12

**Supplementary Table 13 Contribution of right temporal activation to aphasia outcome, above and beyond structural damage**

| | Estimate | SE | *t* | *P* |
|---|---|---|---|---|
| (Intercept) | -6.6512 | 2.2653 | -2.9361 | 0.0047063 |
| Age | -0.023791 | 0.015762 | -1.5094 | 0.13643 |
| SexF | 0.47181 | 0.40264 | 1.1718 | 0.24591 |
| HandLA | -0.315 | 0.5916 | -0.53245 | 0.59638 |
| Educ | 0.068972 | 0.076143 | 0.90583 | 0.36865 |
| StrHem | -0.2336 | 0.68528 | -0.34089 | 0.73438 |
| LogTPO | 2.0877 | 0.5697 | 3.6645 | 0.00052652 |
| PredQAB | 1.7412 | 0.24247 | 7.1808 | 1.2292e-09 |
| LogTPOxPredQAB | -0.26603 | 0.075469 | -3.525 | 0.00081768 |
| R_Temporal | 6.1781 | 2.2519 | 2.7435 | 0.008004 |

Linear regression model: QAB ~ 1 + Age + SexF + HandLA + Educ + StrHem + LogTPO + PredQAB + LogTPOxPredQAB + R_Temporal
Number of observations: 70, Error degrees of freedom: 60
Root Mean Squared Error: 1.54
R-squared: 0.702, Adjusted R-Squared: 0.657
F-statistic vs. constant model: 15.7, p-value = 8.45e-13

**Supplementary Table 14 Contribution of right frontal activation to aphasia outcome, above and beyond structural damage**

| | Estimate | SE | *t* | *P* |
|---|---|---|---|---|
| (Intercept) | -7.1112 | 2.4516 | -2.9007 | 0.005198 |
| Age | -0.020749 | 0.016929 | -1.2257 | 0.22511 |
| SexF | 0.51746 | 0.45083 | 1.1478 | 0.25561 |
| HandLA | -0.3301 | 0.63731 | -0.51796 | 0.60639 |
| Educ | 0.10419 | 0.07916 | 1.3162 | 0.1931 |
| StrHem | 0.078195 | 0.71116 | 0.10995 | 0.91281 |
| LogTPO | 2.0695 | 0.6021 | 3.4371 | 0.0010739 |
| PredQAB | 1.7733 | 0.25792 | 6.8754 | 4.0783e-09 |
| LogTPOxPredQAB | -0.26751 | 0.080173 | -3.3367 | 0.0014593 |
| R_Frontal | 1.7176 | 1.9437 | 0.88367 | 0.3804 |

Linear regression model: QAB ~ 1 + Age + SexF + HandLA + Educ + StrHem + LogTPO + PredQAB + LogTPOxPredQAB + R_Frontal
Number of observations: 70, Error degrees of freedom: 60
Root Mean Squared Error: 1.63
R-squared: 0.669, Adjusted R-Squared: 0.619
F-statistic vs. constant model: 13.5, p-value = 1.69e-11

**Supplementary Table 15 Contribution of multivariable functional activation to aphasia outcome, above and beyond structural damage**

| | Estimate | SE | *t* | *P* |
|---|---|---|---|---|
| (Intercept) | -7.3309 | 2.0508 | -3.5746 | 0.00071468 |
| Age | -0.014668 | 0.014384 | -1.0198 | 0.31208 |
| SexF | 0.37823 | 0.37149 | 1.0181 | 0.31284 |
| HandLA | -0.31723 | 0.53195 | -0.59635 | 0.55326 |
| Educ | 0.097719 | 0.068801 | 1.4203 | 0.16087 |
| StrHem | 0.41017 | 0.63612 | 0.64481 | 0.52159 |
| LogTPO | 2.0943 | 0.51174 | 4.0925 | 0.00013359 |
| PredQAB | 1.5641 | 0.22215 | 7.041 | 2.5017e-09 |
| LogTPOxPredQAB | -0.27667 | 0.067843 | -4.0781 | 0.00014017 |
| L_Temporal | 3.7535 | 1.2526 | 2.9966 | 0.0040124 |
| R_Temporal | 4.4921 | 2.0833 | 2.1562 | 0.035224 |
| L_Frontal | 0.80067 | 0.79449 | 1.0078 | 0.31775 |

Linear regression model: QAB ~ 1 + Age + SexF + HandLA + Educ + StrHem + LogTPO + PredQAB + LogTPOxPredQAB + L_Temporal + R_Temporal + L_Frontal
Number of observations: 70, Error degrees of freedom: 58
Root Mean Squared Error: 1.39
R-squared: 0.768, Adjusted R-Squared: 0.723
F-statistic vs. constant model: 17.4, p-value = 1.46e-14

**Supplementary Table 16 Model predicting aphasia outcomes from structural damage**

| | Estimate | SE | *t* | *P* |
|---|---|---|---|---|
| (Intercept) | -6.6197 | 2.3834 | -2.7775 | 0.0072683 |
| Age | -0.023625 | 0.016583 | -1.4246 | 0.15936 |
| SexF | 0.66736 | 0.41695 | 1.6006 | 0.11463 |
| HandLA | -0.20828 | 0.6211 | -0.33533 | 0.73852 |
| Educ | 0.10347 | 0.079013 | 1.3095 | 0.19528 |
| StrHem | 0.10447 | 0.70926 | 0.1473 | 0.88338 |
| LogTPO | 2.0252 | 0.59893 | 3.3813 | 0.0012636 |
| PredQAB | 1.7426 | 0.25511 | 6.8308 | 4.5219e-09 |
| LogTPOxPredQAB | -0.25829 | 0.079348 | -3.2552 | 0.0018512 |

Linear regression model: QAB ~ 1 + Age + SexF + HandLA + Educ + StrHem + LogTPO + PredQAB + LogTPOxPredQAB
Number of observations: 70, Error degrees of freedom: 61
Root Mean Squared Error: 1.62
R-squared: 0.665, Adjusted R-Squared: 0.621
F-statistic vs. constant model: 15.1, p-value = 6.01e-12

**Supplementary Figure 1**

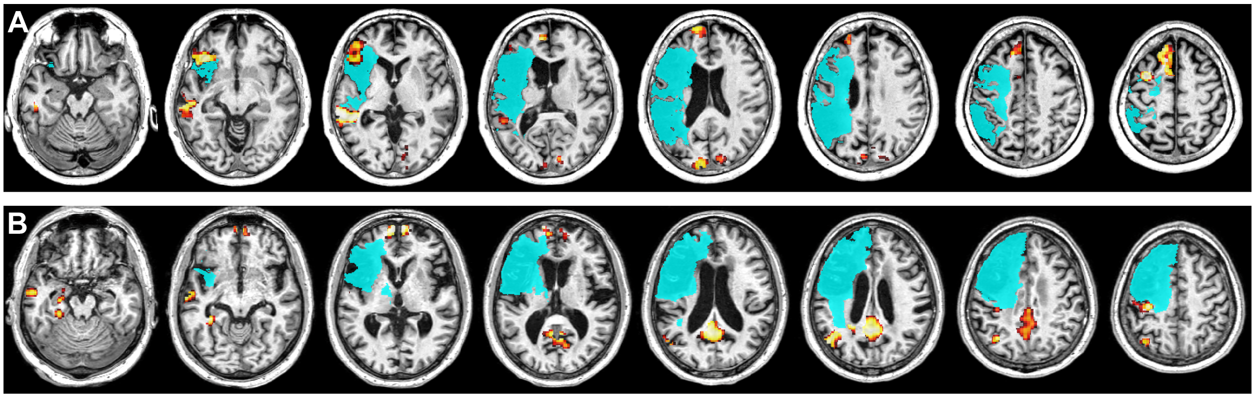


Detailed view of example patient lesions and functional activations. (A) Axial slices showing the lesion and language map for the individual depicted in Figure 4B. (B) Axial slices showing the lesion and language map for the individual depicted in Figure 4C.
